# Irremediable psychiatric suffering in the context of physician assisted death: a Delphi-study

**DOI:** 10.1101/2021.07.19.21260430

**Authors:** Sisco M.P. van Veen, Natalie Evans, Andrea M. Ruissen, Joris Vandenberghe, Aartjan T.F. Beekman, Guy A.M. Widdershoven

## Abstract

**Background:** Patients with a psychiatric disorder (PPD) are eligible to request physician assisted death (PAD) in a small but growing number of jurisdictions, including the Netherlands and Belgium. For this request to be granted, most of these jurisdictions demand that the patient is competent in her request, and that the suffering experienced is unbearable and irremediable. Especially the criterion of irremediability is challenging to establish in patients with psychiatric disorders.

**Aims:** To establish what criteria Dutch and Belgian experts agree to be essential in characterising irremediable psychiatric suffering (IPS) in the context of PAD.

**Method:** A two round Delphi procedure among psychiatrists with relevant experience.

**Results:** Thirteen consensus-criteria were established: five diagnostic and eight treatment-related criteria. Diagnostically, the participants deem a narrative description and attention to contextual and systemic elements necessary. Also, a mandatory second opinion is required. The criteria concerning treatment show that extensive biopsychosocial treatment is needed, and the suffering must be present for several years. Finally, in the case of treatment refusal, the participants agree that there are limits to the number of diagnostic procedures or treatments a patient must undergo.

**Conclusions:** Consensus was found among a Dutch and Belgian expert group on essential criteria for establishing IPS in the context of PAD. These criteria can be used in clinical decision making and can inform future procedural demands and research.

## Introduction

Patients with a psychiatric disorder (PPD) are eligible to request physician assisted death (PAD) in a small but growing number of jurisdictions, including the Netherlands and Belgium. For this request to be granted, most of these jurisdictions demand that the patient is competent in her request, and that the suffering experienced is unbearable and irremediable. (1,2)

The criterion of irremediability is particularly difficult to establish in psychiatric disorders. (3) Retrospective casefile studies of Dutch PPD who died through PAD show if experts disagree on a criterion, it often concerns irremediability. (1,4) A Dutch euthanasia expertise centre recently stated that more clarity on psychiatric irremediability is a priority. (5) A Canadian expert advisory group has also called for more research on the irremediability of psychiatric suffering. (6)

A recent qualitative study on irremediable psychiatric suffering (IPS) among Dutch psychiatrists with experience in establishing IPS in the context of PAD concluded that consensus-criteria are needed to guide current clinical decision-making. (7) Moreover, jurisdictions debating PAD for PPD in the future may benefit from the availability of essential criteria for establishing IPS, developed from the relatively longer practice experiences in the Netherlands and Belgium. If jurisdictions choose to allow PAD for PPD, minimum essential criteria for IPS should also be specified when drafting legislation. Finally, researchers can use criteria of IPS for designing clinical studies on the irremediability of therapy resistant psychiatric disorders and their management.

In this study, we use a Delphi-method to develop expert consensus-criteria for IPS in the context of PAD, focusing on the Netherlands and Belgium where PAD has been permitted in psychiatric practice for over 20 years. Therefore, we address the following research question: what are the criteria that Dutch and Belgian experts agree upon for IPS in the context of PAD for PPD?

## Methods

### Ethical approval, data-management, study design and preregistration

Ethical approval for this study was obtained in the form of a non-WMO declaration from the Medical Ethical Examination Board of Amsterdam UMC / VUmc under registration number D326.

Furthermore, a privacy impact assessment was performed, assuring compliance with European privacy laws. For data-management, the survey tool *Survalyzer* was used, quantitative analysis was performed using *SPSS v25*.*0*, and for qualitative analysis *Maxqda v18*.*2*.*4* was used. Also, the ‘Conducting and REporting of DElphi Studies’ (CREDES) guidelines were followed. (8) The study protocol was preregistered at the Open Science Framework under project code: qx5hy.

### Inclusion criteria

In order to be included in the study (1) participants had to have at least five years of clinical experience as a psychiatrist and (2) they had to have experience with assessing PPD requesting PAD. This could mean that they had investigated a persistent PAD-request from one of their own patients, that they acted as an independent clinical expert in a PAD-procedure, or both. It was not required for them to have actually assisted a patient in dying. No specific exclusion criteria were used.

### Participant selection

First the project group was formed, consisting of the authors of this article, who are Dutch and Belgian experts in PAD in PPD with backgrounds in psychiatry and ethics and different views on PAD for PPD. Through criterion recruitment from the clinical and scientific network of the project group, participants were selected in the Netherlands and Belgium. Diverse perspectives were aimed for by purposely inviting psychiatrists who are known proponents, opponents, or hold a moderate stance on PAD for PPD. Participants in the study were also asked to recommend other experts that met the inclusion criteria *(snowballing)*. An information letter was sent describing that participation will not yield direct benefits and that the main burden is the time investment. Informed consent was obtained from all participants, and everyone gave permission to be acknowledged for their efforts in the final publication, adding to the transparency of the study. Participants were sent an email with a personal link to the online survey. At the beginning of the study, all round one participants were explicitly asked to participate in all subsequent rounds, and during both rounds two reminders were sent.

### Survey design and data analysis

The round one survey was developed during project group meetings using insights from a systematic review and a qualitative interview study among psychiatrists, both on the topic of irremediable psychiatric suffering. (3,7) The criteria were subdivided in three categories: diagnostic criteria, treatment criteria and treatment refusal criteria. First, the participants were asked to give their own definition of IPS. Next, they were asked their opinions on 20 criteria using a 5-point Likert-scale (strongly disagree, disagree, neutral, agree, strongly agree). The participants were encouraged to provide arguments for their ratings in an open comment section. Relevant socio-demographic and professional characteristics were also collected (table 1). Finally, participants were asked whether they would perform PAD for PPD themselves. The survey was validated in a pilot phase, during which three senior psychiatry residents from the Netherlands and Belgium filled out the survey in the presence of the corresponding author using the ‘think aloud’ approach. (9)

**Table 1:**
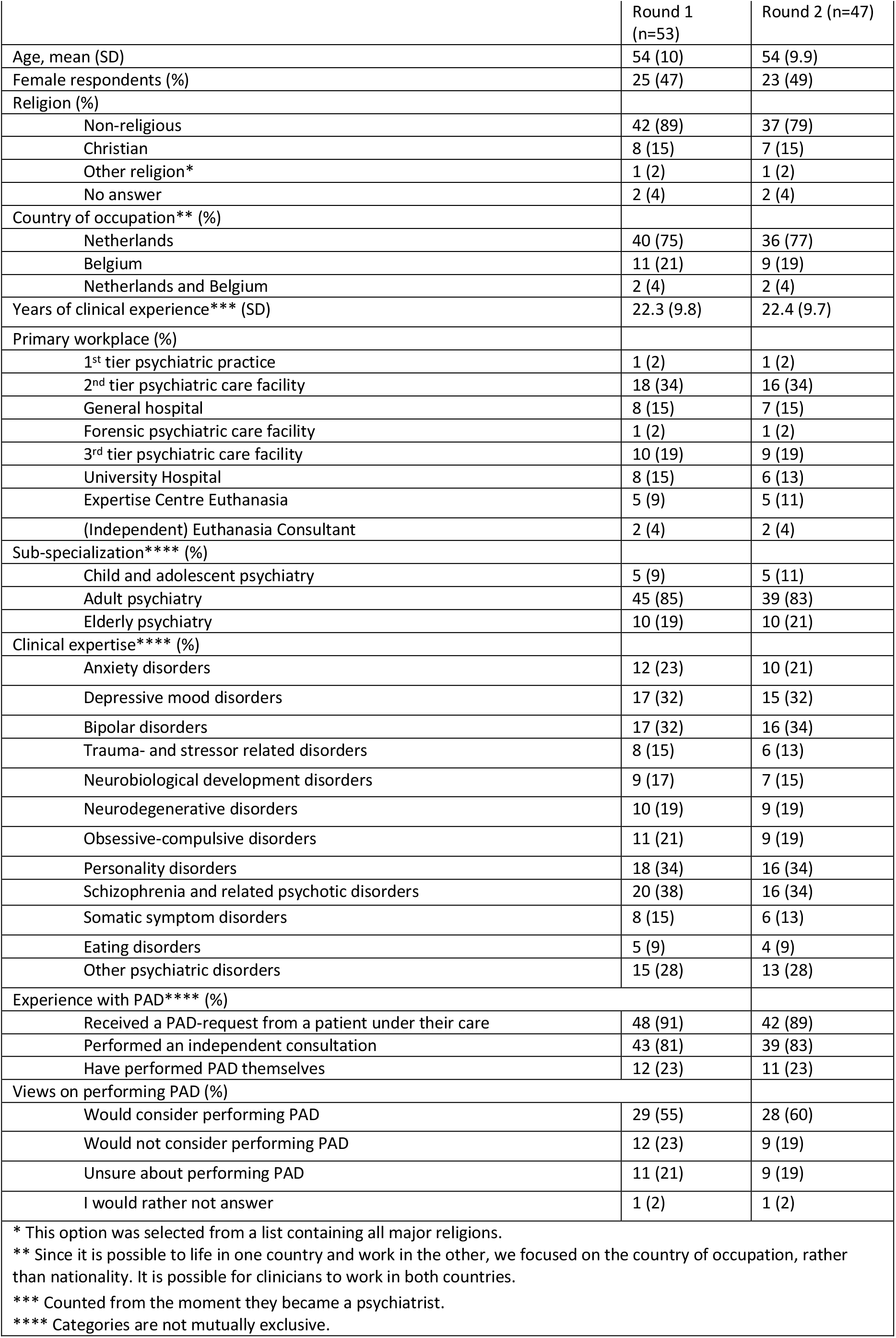
Respondent characteristics

After round one, the open definition and accompanying comments for each criterion were coded and categorized using thematic analysis, paying particular attention to indications that the participant had misunderstood any elements of the criteria or desired more details. (10) The Likert-scales were analysed using basic descriptive statistics. Consensus was defined as 70% of the experts agreeing/disagreeing or strongly agreeing/disagreeing with a statement (i.e. the top or bottom two options on the five-point Likert-scale). (11) The round one results were discussed in two project group meetings and summarized in a feedback report (supplement 1). The open definitions served as inspiration for additional criteria for round two. When the comments showed that criteria were misunderstood or valuable suggestions were given for wording changes, the criteria were modified and included in round two accompanied by a summary of the comments. If the comments lacked relevant arguments and no substantial changes were suggested, the consensus or dissensus about the criterion was accepted. The round two survey was piloted again on one of the senior psychiatric residents using the ‘think aloud’ approach.

The results of round two were discussed in a project group meeting and summarized in a feedback report (supplement 2). Using the same standards as round one it was concluded that the wording was sufficiently clear for all criteria, arguments for agreeing or disagreeing were similar to round one and became repetitive, and no substantial new viewpoints were introduced by participants, indicating response stability. (12) Therefore, the outcomes of round two criteria were accepted and no third round was performed.

## Results

### Participants

67 psychiatrists, meeting the inclusion criteria, responded to an invitation to participate and were sent the survey. 53 psychiatrists completed the first round (79%), of these 47 completed round two (89% of those responding to round 1). Demographic and professional characteristics of participants are shown in Table 1. Of the initial 53 participants, the mean age was 54 and 47% were female. Of these participants, 75% worked in the Netherlands, 21% in Belgium and 4% in both countries. 91% had received a PAD-request from one or more of their own patients, 81% had performed an independent consultation, and 23% had actually performed PAD due to psychiatric suffering.

### Criteria

After two Delphi rounds, consensus was reached for 13 criteria, which can be subdivided into 5 diagnostic and 8 treatment-related criteria (table 2). Below we summarize the iterative process resulting in the consensus-criteria.

**Table 2:**
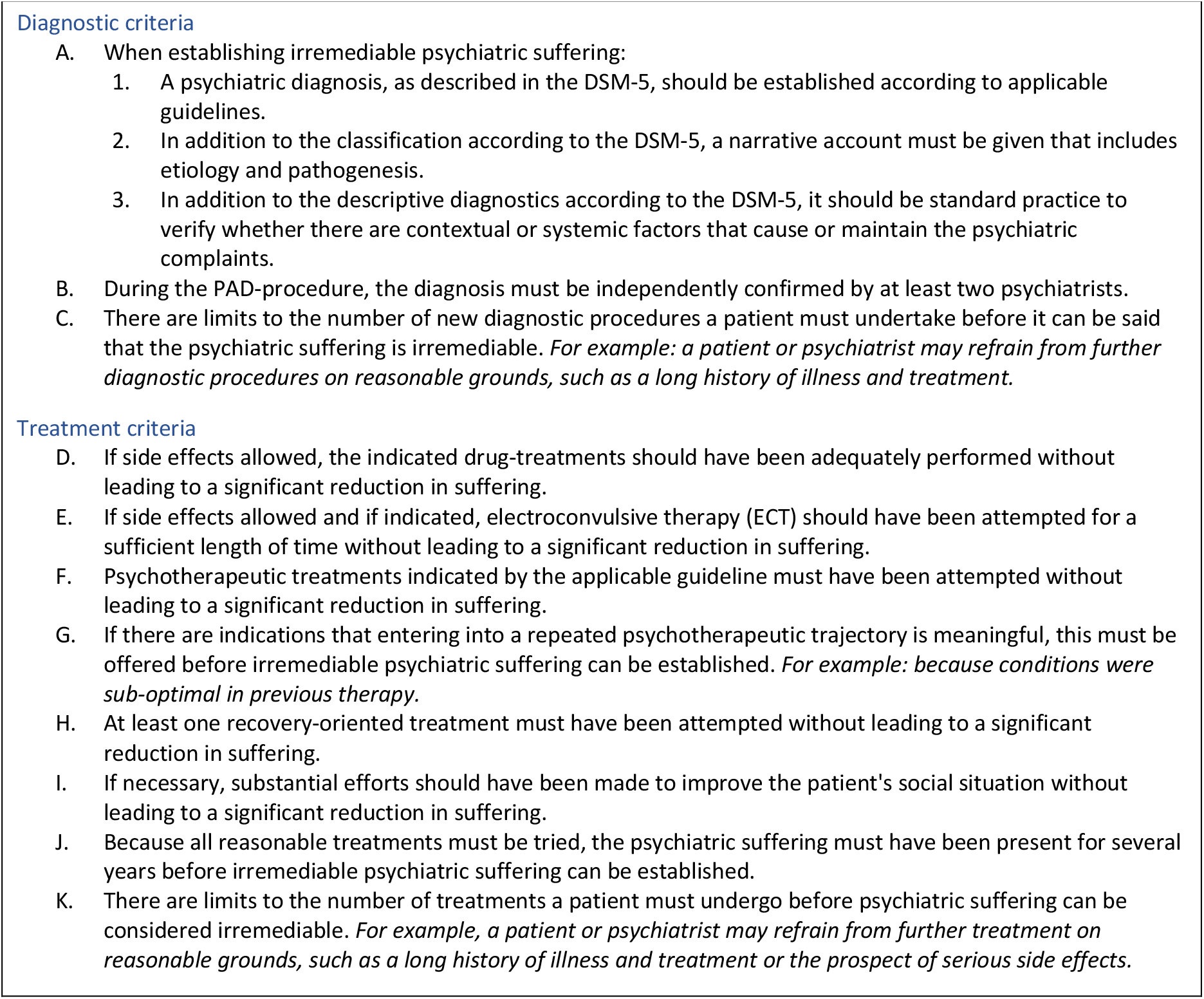
Consensus criteria for irremediable psychiatric suffering in the context of physician assisted death

#### Open definition

At the beginning of round one the participants were asked to give an open definition of IPS in the context of PAD. 52 participants gave detailed definitions. Through thematic analysis, recurrent themes were identified which are summarized below. A full report of the qualitative analysis can be found in supplement one.

Most participants’ definitions specified that a psychiatric disorder should cause suffering which is *persistent, long lasting, chronic or constant*. Also, almost all definitions contained a criterion that the prognosis should be poor, or as one participant defines it:

> *“[IPS is] severe suffering that stems from a psychiatric disorder and cannot be alleviated by any available treatment options*.*” - P16, Dutch, age 50, has experience with PAD as an independent expert*.

Various participants added that extensive treatment must have been tried and failed. Several emphasized the importance of ‘finishing the treatment-protocol’ or ‘trying all evidence-based treatments’ without relief of suffering. Others explicated that only reasonable treatment options can be demanded from the patient. One participant captured both of these perspectives, stating:

> *“Subjectively severe suffering linked to one or more psychiatric diagnoses for which the various treatment options advised by guidelines and accepted within reasonable limits by the patient have been exhausted*.*”-P31, Belgian, 60, has experience with PAD as an independent expert*.

The themes ‘persistence of suffering’, ‘poor prognosis’ and ‘failed treatment’ led to two new criteria for round two (see section on *round two criteria* below).

#### Round one criteria

Round one was subdivided in diagnostic criteria, treatment-related criteria and treatment refusal criteria (table 3). Eight of 20 criteria reached consensus, three of which were diagnostic criteria (table 2: A1, A3 and B), five were treatment criteria (table 2: D, E, F, H & I), none of the treatment refusal criteria reached consensus.

**Table 3:**
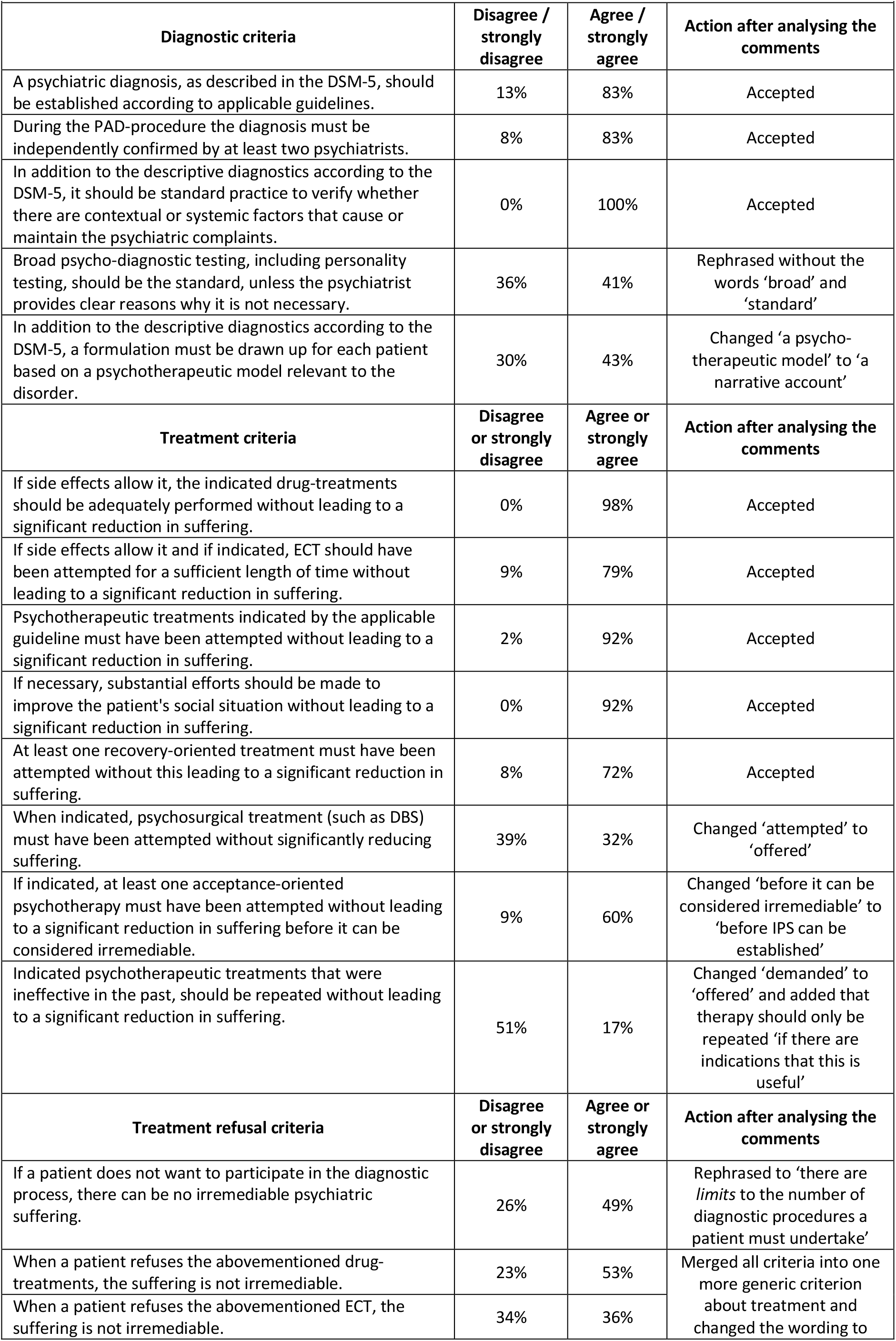
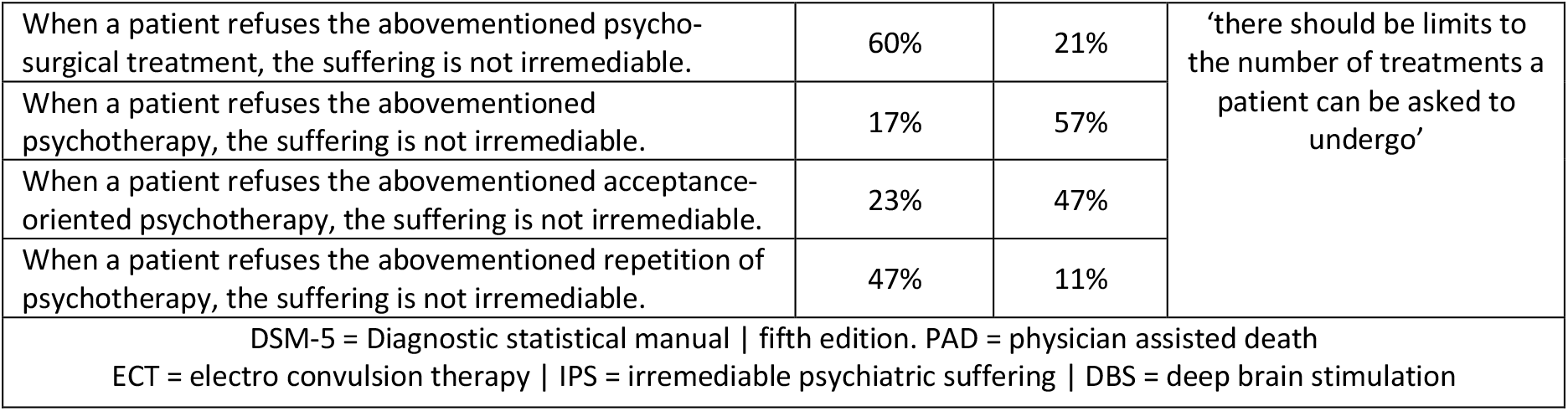
Likert-scale scores of round 1 criteria.

Three of five diagnostic criteria reached consensus in the first round (table 3). The accompanying comments indicated these criteria were sufficiently clear and that no substantial new viewpoints were introduced in the comments. These were not, therefore, repeated in round two. Two diagnostic criteria that did not reach consensus were included in round two after rephrasing guided by the comments (tables 3 and 4).

**Table 4:**
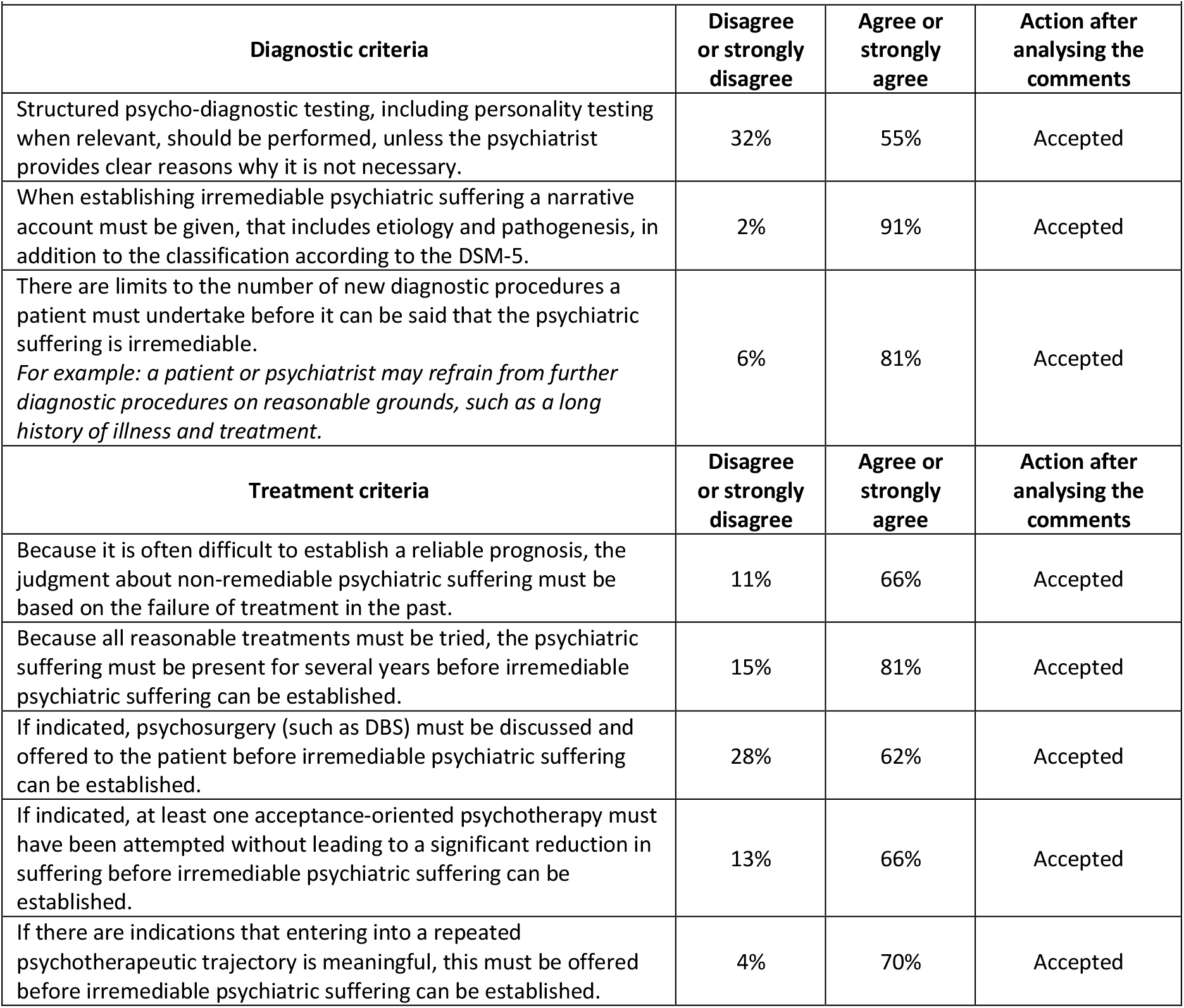
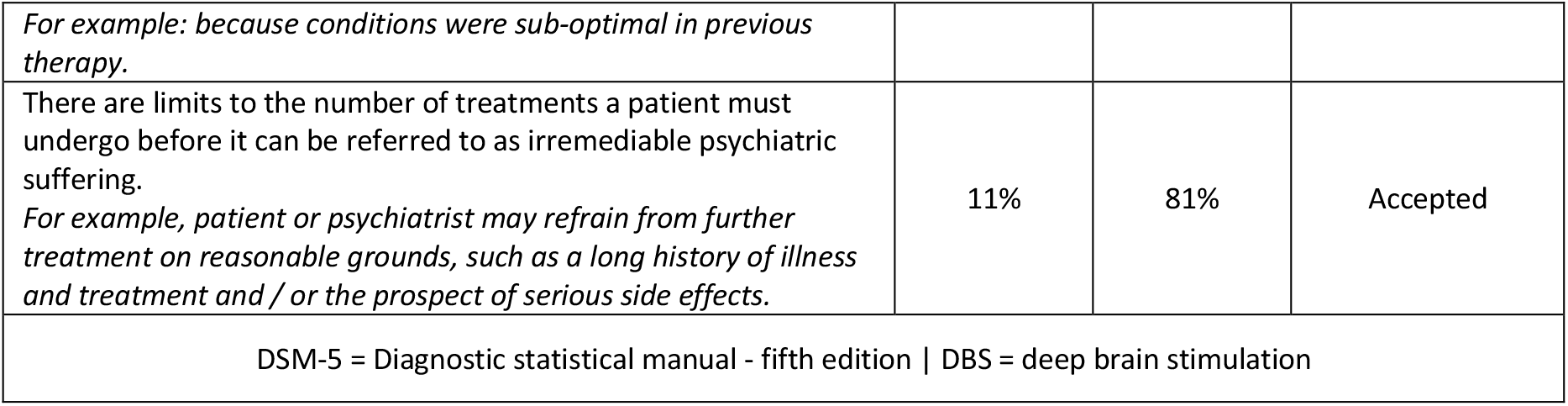
Likert-scale scores of round 2 criteria.

| **Table 4:** Likert-scale scores of round 2 criteria. |  |  |  |
| --- | --- | --- | --- |
| **Diagnostic criteria** | **Disagree or strongly disagree** | **Agree or strongly agree** | **Action after analysing the comments** |
| Structured psycho-diagnostic testing, including personality testing when relevant, should be performed, unless the psychiatrist provides clear reasons why it is not necessary. | 32% | 55% | Accepted |
| When establishing irremediable psychiatric suffering a narrative account must be given, that includes etiology and pathogenesis, in addition to the classification according to the DSM-5. | 2% | 91% | Accepted |
| There are limits to the number of new diagnostic procedures a patient must undertake before it can be said that the psychiatric suffering is irremediable.<br>*For example: a patient or psychiatrist may refrain from further diagnostic procedures on reasonable grounds, such as a long history of illness and treatment.* | 6% | 81% | Accepted |
| **Treatment criteria** | **Disagree or strongly disagree** | **Agree or strongly agree** | **Action after analysing the comments** |
| Because it is often difficult to establish a reliable prognosis, the judgment about non-remediable psychiatric suffering must be based on the failure of treatment in the past. | 11% | 66% | Accepted |
| Because all reasonable treatments must be tried, the psychiatric suffering must be present for several years before irremediable psychiatric suffering can be established. | 15% | 81% | Accepted |
| If indicated, psychosurgery (such as DBS) must be discussed and offered to the patient before irremediable psychiatric suffering can be established. | 28% | 62% | Accepted |
| If indicated, at least one acceptance-oriented psychotherapy must have been attempted without leading to a significant reduction in suffering before irremediable psychiatric suffering can be established. | 13% | 66% | Accepted |
| If there are indications that entering into a repeated psychotherapeutic trajectory is meaningful, this must be offered before irremediable psychiatric suffering can be established. | 4% | 70% | Accepted |
| *For example: because conditions were sub-optimal in previous therapy.* |  |  |  |
| There are limits to the number of treatments a patient must undergo before it can be referred to as irremediable psychiatric suffering.<br>*For example, patient or psychiatrist may refrain from further treatment on reasonable grounds, such as a long history of illness and treatment and / or the prospect of serious side effects.* | 11% | 81% | Accepted |
| DSM-5 = Diagnostic statistical manual - fifth edition \| DBS = deep brain stimulation |  |  |  |

Of the eight initial treatment criteria, five reached consensus in the first round (table 3). From the comments it was clear that all criteria were understood and these were not repeated in round two. Three other criteria did not reach consensus and were adapted based on participants’ comments and repeated in round two (tables 3 and 4).

None of the treatment refusal criteria reached consensus. This appeared to be due to the formulation of the criteria: many participants commented that it is certainly possible that the suffering is irremediable when the patient does not cooperate, but that the irremediability cannot be established in this case. The criteria were reformulated in round two (table 4).

#### Round two criteria

The second round contained nine criteria (table 4). The open definition in round one inspired two new criteria in round two. Of these, one reached consensus (table 2: criterion J). Two diagnostic criteria reached consensus (table 2: criteria A2 and C). Out of three criteria concerning treatment, two reached consensus (table 2: criteria G and K). The comments showed that all criteria were well understood and therefore both the dissensus and the consensus that was found was regarded as valid and no new round was started.

## Discussion

Through a modified Delphi-method, 13 expert consensus-criteria for IPS in the context of PAD were identified, regarding diagnosis and treatment.

### Diagnostic criteria

Participants considered a DSM classification necessary when determining IPS, but even more participants considered the presence of a narrative description and attention to the contextual and systemic elements necessary. These criteria are in line with clinical guidelines describing best practice in psychiatric evaluation. (13,14) The criteria imply that a certain degree of individual interpretation will always be part of the decision-making process concerning IPS in the context of PAD. Further deliberation should focus on what levels of individual interpretation are justifiable. (15)

All participants agreed that a mandatory second opinion should be a criterion of IPS in the context of PAD. This endorses the current due diligence procedures in the Netherlands and Belgium. (16–18) Furthermore, a second opinion can mitigate the risk of interpretative differences. Policymakers from other jurisdictions should take this insight into account when developing their respective due diligence procedures for psychiatric PAD.

The participants also agree that, although there must be evidence of a substantial clinical history (described in detail below), there should be limits to the number of diagnostic procedures a patient has to undergo before IPS can be established. Especially when this patient has a long history of illness and treatment. This criterion suggests that if the patient refuses certain diagnostic procedures, it does not automatically mean that IPS cannot be established. It may be justified to only demand additional diagnostic procedures, such as neuropsychological or personality testing, when there is a reasonable chance that this will lead to new treatment options.

### Treatment criteria

The participants agree that substantial treatments have to have failed before IPS can be established. This is in line with earlier studies. (3,7,19)

The participants also agree that, because all reasonable treatments must be tried, the suffering should be present for several years. This criterion reflects the notion that persistence of suffering is not only time dependent but also treatment dependent. (20) Implementation of this criterion as a due diligence requirement might provide clarity for patients and regulators about the high threshold of irremediability in the context of PAD for PPD. It may also be relevant for distinguishing PAD-requests from impulsive suicidality. (21)

The results also show that experts take both biological and psychological treatments, and social interventions, into account when establishing IPS. This is in line with the biopsychosocial model of psychiatric suffering and treatment, which was introduced by George Engel in 1977, and is still highly influential in contemporary psychiatry. (22) In the context of PAD it can serve as a helpful framework to assess individual treatment criteria.

Regarding the biological treatments, there is consensus that medication and ECT should have been tried, but not psychosurgery. Based on the comments, two main reasons for dissensus emerge: participants find psychosurgery too experimental, too invasive, or both. This shows that effectiveness and proportionality should be taken into account when deciding on treatment criteria.

Participants also agree that appropriate psychotherapeutic treatments must have failed before IPS can be established. Psychiatrists especially supported repetition of psychotherapy if there are indications that earlier therapy was ‘performed inadequately’. This criterion should be used with caution, as it is difficult to reliably evaluate the quality of earlier psychotherapy, and knowledge about the efficacy of repeated psychotherapy for therapy-resistant psychiatric complaints is lacking. (7,23) There was no consensus regarding the criterion that acceptance-based therapy should be attempted. This is at odds with the suggestion that working towards acceptance of suffering can be seen as a subsidiary option to psychiatric PAD. (3)

The participants consider social interventions, including recovery-oriented approaches, important in the context of PAD. Also, if necessary, substantial efforts should be made to improve the patient’s social situation. This may be read as support for the often use d argument against PAD stating that when a patient with a psychiatric disorder wants to die, we should improve their situation, not offer PAD. (24) But the criterion also implies that when there is no need for improving social support or if proper support does not reduce suffering, IPS may still be established and PAD may still be justified.

Finally, there was consensus that there should be limits to the number of treatments a patient has to undergo before IPS can be established. This allows for treatment refusal, which is an important theme in the debate about IPS in the context of PAD. (3) However, as none of the specific treatment refusal criteria reached consensus, no conclusions can be drawn regarding what specific psychiatric treatments the patient should undergo before IPS can be established in the context of PAD. The current criterion, that only states that there should be *limits*, leaves room for interpretive differences between psychiatrists, patients and other stakeholders. This can be seen as an argument for a diligent assessment of IPS, which requires experience and expertise of participants, as well as joint deliberation, in order to apply the criterion to an individual case.

### Strengths and weaknesses

A strength of this study is that we were able to access a substantial group of psychiatrists with ample experience in assessing PAD-requests, and representing different views on psychiatric PAD.

A limitation of a consensus-building Delphi survey is that the structure of the questionnaire is determined by the researchers, and participants’ comments are interpreted by the project group, limiting the influence of the participants. Moreover, no widely accepted benchmark exists within the research community of what constitutes an adequate level of consensus. Also, because we had to change the criteria in between rounds we were not able to calculate response-stability for any of our questions, which could have been a valuable addition to the current qualitative analysis of response stability. (12)

### Recommendations for practice and research

We recommend implementing these consensus criteria in the due diligence procedures in the Netherlands and Belgium, in order to contribute to more uniformity and fewer fundamental disagreements when assessing the irremediability of psychiatric suffering in the context of PAD. The criteria should be seen as essential, but not necessarily sufficient, and have to be applied with due expertise.

On a wider scale, we recommend that other jurisdictions which allow PAD for PPD, or are currently discussing options for doing so, will consider the importance of specifying minimum and essential criteria for the establishment or IPS. We hope that the criteria agreed on by Dutch and Belgian experts can be used to inform the development of guidance in other jurisdictions. We recommend that this study is replicated in other countries to see whether similar criteria are agreed upon. We also recommend to explore patient views on irremediability more deeply.

Although the criteria are specifically established for patients requesting PAD, they can serve as inspiration for examining psychiatric irremediability in general. Psychiatry is beginning to understand how to rationally deploy sequential treatment modalities, but when to stop treatment is rarely discussed. This is problematic, because continuing to treat patients if no effect is to be expected could be harmful, especially when coercion and burdensome procedures are used. By reliably establishing that suffering is irremediable, we may recognize further curative treatment as futile and shift to palliative approaches. (25,26)

### With their permission, we want to acknowledge all participants

In alphabetical order: T. van Balkom, A. Batalla, A. Been, A. ten Berg, T. Birkenhäger, D. Bloemkolk, M. van den Bergh, A. Bosmans, L. van Bouwel, G. de Cuypere, L. Dil, A. Dols, R. van Duursen, J. Garcia Barnet, G. Glas, K. Goethals, D. van Grootheest, R. Harbers, O. van den Heuvel, M. de Hert, C. Hoff, J. Hovens, C. Huyser, R. Keet, I. Klijn, F. van Koningsveld, P. Kölling, J. Lenssen, J. Luykx, T. Ingenhoven, P. Naarding, H. van Nuffel, P. Neuteleers, D. Peeters, Y. Roke, R. Rotteveel, L. Tak, E. Thys, P. Schulte, G. Smid, R. Snoeij, E. Spuijbroek, M. Soons, P. Stärcke, G. Steegen, J. Steenmeijer, B. Vandekerkhove, C. Vanmechelen, R. de Veen, I. van der Velden, N. Vulink, J. van Waarde & D. de Wachter.

## Data Availability

The data referred to in the manuscript is available through osf.io/qx5hy/

