## Supplement 1 for "Irremediable psychiatric suffering in the context of physician assisted death: a Delphi-study"

### Irremediable psychiatric suffering - Delphi round 1 - Feedback report

#### Table of contents

Table of contents 1

Executive summary 2

Demographics 3

- Respondent selection and response rate 3
- Respondent characteristics 3
- Work related characteristics 3
- Experience with PAD 4

Quantitative data summary 5

- Consensus criteria 5
- Dissensus criteria 6

Qualitative content analysis of respondent comments 8

- Open definition 8
- Consensus criteria 11
- Dissensus criteria 17

Hyperlink to complete results (Dutch) 28

### Executive summary

The aim of this Delphi-study is to find expert-consensus on the concept of irremediable psychiatric suffering (IPS) in the context of physician assisted death (PAD). The first round contained two elements. First, we asked 67 psychiatrists with experience in PAD-requests to give a definition of IPS in the context of PAD. Second, we asked them to rate provided criteria on Likert scales and comment on their choices. The response rate of the first round was 79% (53/67). 77% of respondents worked in the Netherlands, 23% in Belgium. 91% received a PAD request from a patient in their care, 77% performed an independent clinical consultation during a PAD-procedure, 10 respondents had performed an independent *procedural* consultation (functioning as a SCEN- or LEIF-physician). 22.6% performed PAD themselves.

The open definition was analyzed using a qualitative content analysis. Three categories emerged from the respondents' definitions: diagnostic factors, treatment factors and prognostic factors. Many respondents included a demand that a psychiatric disorder should be present in their definition. Many also stated that extensive treatment should be tried, though respondents conceptualized this differently. Another common element in the definitions was that the prognosis must be poor.

The provided criteria were subdivided in diagnostic, treatment and criteria concerning refusal. Three of five diagnostic criteria reached consensus. First, a psychiatric diagnosis should be present. Second, this diagnosis should be confirmed by an independent psychiatrist. Third, it should be verified whether contextual or systemic factors cause or maintain the psychiatric complaints. Two other diagnostic criteria did not reach consensus. First, whether broad psychodiagnostics testing should be standard. Second, whether in addition to the descriptive diagnosis a psychotherapeutic formulation from a relevant model should be drawn up.

Of the eight treatment criteria, five reached consensus. First, indicated drug-treatment should be adequately performed if side-effects allow it. Second, electro convulsion treatment must be tried if indicated. Third, indicated psychotherapy must have been attempted. Fourth, substantial efforts should be made to improve the patient's social situation. Fifth, at least one recovery-oriented treatment must have been attempted. Three criteria did not reach consensus. First, dissensus existed whether psychosurgical treatment must have been attempted if indicated. Second, whether acceptance-oriented psychotherapy must have been attempted. And third, whether indicated psychotherapeutic treatments should be repeated.

Finally, we asked whether the suffering could still be irremediable if the patient refused any of the abovementioned treatments or diagnostic procedures. None of these criteria reached consensus. This may be due to the fact the question was internally inconsistent (which many respondents noticed): suffering can be irremediable when the patient does not cooperate, but this irremediability cannot be established. A relevant factor seems to be what grounds the patient has for refusal; if these grounds are reasonable (for instance because of a long history of treatments and potential side-effects), this may be a reason for considering psychiatric suffering as irremediable.

We used the definitions, Likert-scales and comments to design the second Delphi-round. The criteria that reached consensus will not be repeated.

### Demographics

#### Respondent selection and response rate

We purposively sampled potential respondents from the authors' respective networks, resulting in 67 psychiatrists fitting the inclusion criteria and showing interest in participating. The survey was sent on May 27th, 2020, and remained open for four weeks, two reminders were sent. 53 of the 67 experts completed the survey (79%).

#### Respondent characteristics

The age of respondents ranged from 34 to 73, with a mean of 53,6 (SD 10,0). 53% of the respondents were male, 47% were female.

79.2% of respondents identified as non-religious, 17% identified as religious, and 4% of respondents declined to answer. Of the respondents that identified as religious, 8 respondents (89%) identified as Christian, 1 respondent (11%) choose the category 'other religion' from a list that contained all major religions.

#### Work related characteristics

41 (77%) respondents worked in the Netherlands, 12 (23%) respondents worked in Belgium. Their clinical experience ranged from 5 to 40 years, with a mean of 22.3 years (SD 9,8).

34% of respondents primarily worked in a (2nd tier) psychiatric care facility, 15% worked in a general hospital, 15% worked in an university hospital, 10% worked in a specialized (3rd tier) psychiatric care facility, 9% worked at the Euthanasia Expertise Centre, 4% primarily worked as a euthanasia consultation specialist (called SCEN-physician in Netherlands or LEIF-physician in Belgium). 1 respondent worked as an independent (1st tier) psychiatrist (2%), and 1 respondent worked as a forensic psychiatrist (2%).

9% of respondents were specialized in child- and adolescent psychiatry, 85% in adult psychiatry and 19% in elderly psychiatry (categories where not mutually exclusive).

When asked about their areas of expertise (also not mutually exclusive), 17 respondents (32%) identified as expert on depressive mood disorders, 12 (23%) as expert on anxiety disorders, 17 (32%) on bipolar disorders, 8 (15%) on trauma- and stressor related disorders, 9 (17%) on neurobiological development disorders, 10 (19%) on neurodegenerative disorders, 11 (21%) on obsessive-compulsive disorders, 18 (34%) on personality disorders, 20 (38%) on schizophrenia and related psychotic disorders, 8 (15%) on somatic symptom disorders, 5 (9%) on eating disorders and 15 (28.3%) on other psychiatric disorders.

#### Experience with PAD

48 (91%) of the respondents received a PAD-request from a patient that was under their care. 41 (77%) respondents have performed an independent *clinical* consultation for a patient with a PAD request. 10 (19%) respondents had performed an independent *procedural* consultation for a patient with a PAD request (functioning as a SCEN- or LEIF-physician). 12 (23%) respondents had performed PAD themselves. The number of PADs performed by a single respondent ranged from 1 (N=7) to 89 (N=1), with a median of 1.

55% of respondents would consider performing PAD themselves at one point, 23% of respondents would not, and 21% was not sure about this.

### Quantitative data summary

#### Consensus criteria

| Diagnostic criteria | % agree or strongly agree |
|---|---|
| A psychiatric diagnosis, as described in the DSM-5, should be established according to applicable guidelines. | 83% |
| During the PAD-procedure the diagnosis must be independently confirmed by at least two psychiatrists. | 83% |
| In addition to the descriptive diagnostics according to the DSM-5, it should be standard practice to verify whether there are contextual or systemic factors that cause or maintain the psychiatric complaints. | 100% |
| **Treatment criteria** | **% agree or strongly agree** |
| If side effects allow it, the indicated drug-treatments should be adequately performed without leading to a significant reduction in suffering. | 98% |
| If side effects allow it and if indicated, electroconvulsive therapy (ECT) should have been attempted for a sufficient length of time without leading to a significant reduction in suffering. | 79% |
| Psychotherapeutic treatments indicated by the applicable guideline must have been attempted without leading to a significant reduction in suffering. | 92% |
| If necessary, substantial efforts should be made to improve the patient's social situation without leading to a significant reduction in suffering. | 92% |
| At least one recovery-oriented treatment must have been attempted without this leading to a significant reduction in suffering. | 72% |

#### Dissensus criteria

| Diagnostic criteria | % disagree or strongly disagree | % agree or strongly agree |
|---|---|---|
| Broad psycho-diagnostic testing, including personality testing, should be the standard, unless the psychiatrist provides clear reasons why it is not necessary. | 36% | 41% |
| In addition to the descriptive diagnostics according to the DSM-5, a formulation must be drawn up for each patient based on a psychotherapeutic model relevant to the disorder. | 30% | 43% |
| **Treatment criteria** | **% disagree or strongly disagree** | **% agree or strongly agree** |
| When indicated, psychosurgical treatment (such as Deep Brain Stimulation) must have been attempted without significantly reducing suffering. | 39% | 32% |
| If indicated, at least one acceptance-oriented psychotherapy must have been attempted without leading to a significant reduction in suffering before it can be considered irremediable. | 9% | 60% |
| Indicated psychotherapeutic treatments that were ineffective in the past, should be repeated without leading to a significant reduction in suffering. | 51% | 17% |
| **Criteria concerning refusal** | **% disagree or strongly disagree** | **% agree or strongly agree** |
| If a patient does not want to participate in the diagnostic process, there can be no irremediable psychiatric suffering. | 26% | 49% |
| When a patient refuses the abovementioned drug-treatments, the suffering is not irremediable. | 23% | 53% |
| When a patient refuses the abovementioned ECT, the suffering is not irremediable. | 34% | 36% |

| | | |
|---|---|---|
| When a patient refuses the abovementioned psycho-surgical treatment, the suffering is not irremediable. | 60% | 21% |
| When a patient refuses the abovementioned psychotherapy, the suffering is not irremediable. | 17% | 57% |
| When a patient refuses the abovementioned acceptance-oriented psychotherapy, the suffering is not irremediable. | 23% | 47% |
| When a patient refuses the abovementioned repetition of psychotherapy, the suffering is not irremediable. | 47% | 11% |

### Qualitative content analysis of respondent comments

#### Open definition

52 respondents composed their own definition of irremediable psychiatric suffering. The definitions were varied, but several elements came back regularly: diagnostic factors, treatment factors, and prognostic factors.

| Category label | Codes | Example quotes | Steering group response |
|---|---|---|---|
| Diagnostic factors | 1. Psychiatric disorder present | *"feelings, thoughts, and behaviors arising from a psychiatric disorder"* | Code 1 has already been addressed in questionnaire 1.<br>Codes 2-5 are related to the unbearableness of suffering, not to the irremediability. In the steering group we discussed that although unbearableness is an important and interesting concept in the context of PAD, it is beyond the scope of this study. Therefore, we will not explore this concept any further.<br>We do however realize that this might be confusing for the respondents and added the following explanation to the introduction of survey 2 [in Dutch]: *'Several comments in round 1 addressed the unbearability of suffering. This is an important concept in the context of euthanasia and in practice intolerability and irremediability will often go hand in hand. But because our current goal is to arrive at clear criteria for irremediable psychological suffering, we would like to ask you to leave the intensity/unbearability of suffering aside for now.'* |
| | 2. Suffering must be unbearable | *"Prolonged and severe psychological suffering that causes unbearable suffering for the person concerned (…)"* | |
| | 3. Quality of life must be impaired | *"which has seriously affected the quality of life* | |
| | 4. Suffering is subjective | *"Subjectively very severe suffering linked to one or more psychiatric diagnoses"* | |
| | 5. Suffering must be palpable | *"For me, long-term suffering must be present that is palpable for me."* | |
| Treatment factors | 6. Treatment must have been ineffective | *"Having psychological complaints and symptoms for which treatments do not provide relief"* | Code 6, 7, 8, 10-12 have already been addressed in questionnaire 1. |

| | | | |
|---|---|---|---|
| | 7. The protocol must be ‘finished’ | *“suffering where the treatments according to the guideline have not helped”* | Code 9: The specifier ‘reasonability’ is used in the Dutch PAD-law, but we did not use it in round 1. The concept of reasonability will be addressed in round 2 through rephrasing of the ‘refusal’ criteria. |
| | 8. Evidence based treatment options must have been tried | *“Psychiatric suffering for which no substantial improvement or relief can be expected through evidence-based treatment”* | |
| | 9. Reasonable treatment options must have been tried | *“Reasonable treatment options must have been exhausted”* | |
| | 10. Recovery based treatment must have been tried | *“Social psychiatric treatments aimed at recovery are not feasible or do not provide relief from the complaints.”* | |
| | 11. Acceptance based treatment must have been tried | *“(…) where even treatment aimed at acceptance does lead to improvement.”* | |
| | 12. Biopsychosocial treatment must have been tried | *“(…) that persists after - at least all regular - treatments with a bio-psycho-social approach (…)”* | |
| Prognostic factors | 13. The suffering must be persistent | *“Psychiatric suffering, which is very likely to remain - almost - unchanged over the course of years.”* | The concepts addressed in codes 13-15 were mentioned in the open definition, but are not explicitly mentioned in any of the criteria in questionnaire 1.<br><br>In the steering group discussion, we decided to add the following two criteria to round 2. |
| | 14. The suffering must be irreversible | *“Suffering that is irreversible”* | |

| | | | |
|---|---|---|---|
| | 15. Perspective and hope must be absent | *"No hope and expectation anymore for improvement."* | New criterium 1:<br><br>**'Because it is often difficult to establish a reliable prognosis, the judgment about non-remediable psychiatric suffering must be based on the failure of treatment in the past.'**<br><br>In Dutch: **'Omdat het stellen van een betrouwbare prognose vaak moeilijk is moet het oordeel over niet remedieerbaar psychiatrisch lijden gebaseerd worden op het niet slagen van behandeling in het verleden.'**<br><br>Rationale for adding this criterium: several respondents use prognostic elements for their definition of irremediability (prospective), others base their definition more on exhausted treatment factors (retrospective).<br><br>This is interesting because accurate prognostics is notoriously difficult in psychiatry. Some see this is as an important reason for excluding psychiatric patients from PAD, for instance by experts in Canada. Other see this as unjustified for we also know that there are patients that will not recover, their suffering is truly irremediable.<br><br>A reasonable solution to this dilemma could be to introduce a retrospective view on irremediability in psychiatry, which is based on the absence of reasonable other solutions for the suffering, instead of a definition based on prognosis. The abovementioned criterium aims to do this.<br><br>New criterium 2:<br><br>**'Because all reasonable treatments must be tried, the psychiatric suffering must be present for several years before irremediability can be established.'** |

| | | | |
|---|---|---|---|
| | | | In Dutch: **'Omdat alle redelijke behandelingen geprobeerd moeten zijn, moet het psychiatrisch lijden enkele jaren aanwezig zijn, voordat niet-remedieerbaarheid vastgesteld kan worden.'**<br><br>Rationale for adding this criterium: this builds on the abovementioned criterium, if the suffering has been present for several years, the chances of (spontaneous) recovery are smaller than when the suffering is relatively short lived. We discussed using the word 'persistent', but decided against this for this might lead to confusing given the typically waxing-waning course of mental disorders. |

#### Consensus criteria

Consensus was defined that 70% or more of the respondents agreed or strongly agreed, or inversely disagreed or strongly disagreed with a criterium. Eight of the twenty criteria reached consensus in the first round.

| **Diagnostic criteria** | | | |
|---|---|---|---|
| **Criterium** | **Respondents view on importance of the criterium** | **Example quotes** | **Steering group response** |
| 2. A psychiatric diagnosis, as described in the DSM-5, should be established according to applicable guidelines. | There was a broad consensus (83%) for this criterium.<br><br>Respondents saw a psychiatric diagnosis as the basis for treatment and subsequently as a basis for establishing irremediability. | *"a DSM diagnosis by a skilled psychiatrist is a foundation for good treatment."*<br><br>*"As long as the DSM is used as a standard for diagnostics, a DSM-diagnosis must have been made"*<br><br>*"In my opinion, psychological suffering without a psychiatric disorder cannot be a reason for euthanasia"*<br><br>*"There is no necessary connection between a diagnosis and the degree of suffering, whether or not it would be bearable* | Consensus was reached for this criterium.<br><br>From the comments we can conclude that the criterium was sufficiently clear. No exemptions or alternative wording options were proposed by respondents that can be expected to |

| | | | |
|---|---|---|---|
| | Several respondents did however argue that a descriptive diagnosis is more important than a DSM-5 classification and questioned the validity of the DSM. Others saw the DSM as valuable for its reliance and international use.<br><br>A diagnosis was also seen as an important way of separating 'psychiatric' from 'psychological' suffering. Some saw this as an important justification for or against PAD.<br><br>Other respondents remarked that a diagnosis says little to nothing about the individual suffering. | *or hopeless; this requires a personal (N=1) investigation and cannot be captured in a global static formula."* | significantly alter the outcome in the following round.<br><br>We will therefore not repeat this criterium in the 2nd round and accept it as a consensus-criterium for irremediable psychiatric suffering. We will however describe these criteria in the executive summary. |
| 3. During the PAD-procedure the diagnosis must be independently confirmed by at least two psychiatrists. | 83% of respondents agreed with this criterium. The weight of the decision to assist someone in dying appeared to be an important reason for demanding a 2nd opinion. | *"Because the decision is a serious one and has major consequences, there must be maximum clarity about diagnosis and assessment of the seriousness of the suffering"*<br><br>*"No psychiatrist can make such an important decision alone. Certainly, in psychiatry, much is not measurable and can be interpreted differently. (…)"* | Consensus was reached for this criterium.<br><br>From the comments we can conclude that the criterium was sufficiently clear. No exemptions or alternative wording options were proposed by respondents that |

| | | | |
|---|---|---|---|
| | Also the complexity and dynamic nature of psychiatric diagnoses were mentioned as a reason to ask for independent confirmation. Psychiatrists found support from a 2nd opinion that they were on the right path.<br><br>Several respondents mentioned that a different DSM classification was not necessary.<br><br>Psychiatrists that opposed a 2nd opinion argued that there is no necessary link between the diagnosis and suffering. They also found it too burdensome for the patient. | *"I think it is important to confirm a psychiatric diagnosis by a second psychiatrist, the classification may be slightly different."*<br><br>*"There is no necessary link between the diagnosis and the unique suffering of the person with a psychiatric illness."*<br><br>*"I think one psychiatrist is sufficient. You can't expose a patient to too many highly stressful investigators. They experience that as having to take an exam every time."* | can be expected to significantly alter the outcome in the following round.<br><br>We will therefore not repeat this criterium in the 2nd round and accept it as a consensus-criterium for irremediable psychiatric suffering. We will however describe these criteria in the executive summary. |
| 5. In addition to the descriptive diagnostics according to the DSM-5, it should be standard practice to verify whether there are contextual or systemic factors that | All respondents agreed with this statement (100%). Many commented that this is part of the standard diagnostic process, and different respondents mention that psychiatric complaints are always context dependent. | *"This is also part of normal diagnostics, see the psychiatric diagnostics guideline"*<br><br>*"This is so natural. Of course, you should not only look at the patient but also at his context and system around it. A person exists in a social context."* | Consensus was reached for this criterium.<br><br>From the comments we can conclude that the criterium was sufficiently clear. No exemptions or alternative wording options were proposed by respondents that can be expected to |

| | | | |
|---|---|---|---|
| cause or maintain the psychiatric complaints. | | | significantly alter the outcome in the following round.<br><br>We will therefore not repeat this criterium in the 2nd round and accept it as a consensus-criterium for irremediable psychiatric suffering. We will however describe these criteria in the executive summary. |
| | **Treatment criteria** | | |
| **Criterium** | **Respondent views on importance of the criterium** | **Example quotes** | **Steering group response** |
| 8. If side effects allow it, the indicated drug-treatments should be adequately performed without leading to a significant reduction in suffering. | 98% of respondents agreed with this statement.<br><br>1. Drug-treatment is standard practice.<br>2. Psychiatric suffering can be treated with drugs.<br>3. ‘reasonability’ is important when considering whether drug-treatment is indicated.<br>4. One respondent argued that if drugs were discontinuated | *“This seems obvious to me, if a treatment has not been carried out adequately, I cannot be convinced that a condition really cannot be cured.”*<br><br>*“You want to know whether an adequate attempt has been made.”*<br><br>*“Seems clear to me. We must, however, be reasonable here. Some treatments are not feasible at a certain point (…)”*<br><br>*“In addition to this, the medication treatment should also be built up very gradually in the case of someone who suffered from side effects. (…)”* | Consensus was reached for this criterium.<br><br>From the comments we can conclude that the criterium was sufficiently clear. No exemptions or alternative wording options were proposed by respondents that can be expected to significantly alter the outcome in the following round.<br><br>We will therefore not repeat this criterium in the 2nd round and accept it as a consensus-criterium for irremediable |

| | | | |
|---|---|---|---|
| | because of side-effects a rechallenge with a slow build-up should be considered. | | psychiatric suffering. We will however describe these criteria in the executive summary. |
| 10. If side effects allow it and if indicated, electroconvulsive therapy (ECT) should have been attempted for a sufficient length of time without leading to a significant reduction in suffering. | **Agree (79%)**<br><br>- ECT is a safe and effective treatment for different psychiatric disorders.<br><br>- ECT should not be applied 'to meet the PAD-requirements'.<br><br>- Because ECT is invasive, patients should be able to refuse this. | *"ECT is an effective, safe treatment and should always be considered when indicated."*<br><br>*"But limited to the positive ECT indication (and not the last resort, unexplored indications such as mood complaints in autism and personality disorders)"*<br><br>*"Much more often people with a wish for euthanasia have a complex psychiatric clinical picture, of which it can sometimes be estimated in advance that ECT will give little chance of improvement. If the patient does not want it, then the obligation to undergo ECT is debatable."* | Consensus was reached for this criterium.<br><br>From the comments we can conclude that the criterium was sufficiently clear. No exemptions or alternative wording options were proposed by respondents that can be expected to significantly alter the outcome in the following round.<br><br>We will therefore not repeat this criterium in the 2nd round and accept it as a consensus-criterium for irremediable psychiatric suffering. We will however describe these criteria in the executive summary. |
| 14. Psychotherapeutic treatments indicated by the applicable guideline must have been attempted | **Agree (92%)**<br><br>- Psychotherapy is seen as an essential treatment-step for psychiatric suffering. | *"Psychotherapeutic treatment offers a chance to improve the condition or learn to handle the complaints better, which should not be left unused, especially in the case of chronic problems that is of great importance."* | Consensus was reached for this criterium.<br><br>From the comments we can conclude that the criterium |

| | | | |
|---|---|---|---|
| without leading to a significant reduction in suffering. | Respondents further mention that it is evidence based and non-invasive.<br><br>- Further comments again mention the importance of reasonability: not all different psychotherapies have to be tried.<br><br>- And one respondent emphasizes the importance of treatment motivation for the indication of psychotherapy. | *"Only if it is reasonable. There is always a psychotherapeutic treatment that has not yet been tried."*<br><br>*"Psychotherapeutic treatments are particularly likely to be successful in (highly) motivated patients. If the patient's motivation stems exclusively from his or her wish for euthanasia, the indication of psychotherapy (perhaps to the exclusion of very short-term behavioral therapy interventions) is in general not fitting."* | was sufficiently clear. No exemptions or alternative wording options were proposed by respondents that can be expected to significantly alter the outcome in the following round.<br><br>We will therefore not repeat this criterium in the 2nd round and accept it as a consensus-criterium for irremediable psychiatric suffering. We will however describe these criteria in the executive summary. |
| 20. If necessary, substantial efforts should be made to improve the patient's social situation without leading to a significant reduction in suffering. | **Agree (92%)**<br><br>Several respondents emphasized that in their experience social factors played an important role in the death wish of patients with a psychiatric disorder.<br><br>Other respondents elaborated that changing social factors can be challenging, especially for patients with a psychiatric disorders that request PAD. | *"In my opinion, this is underexposed, but social factors (loss of work/role functioning, loneliness) may be one of the main reasons why patients with a psychiatric disorder develop a desire for euthanasia."*<br><br>*"Context and social conditions must have been tried exhaustively to improve. Euthanasia should not be used because of the social circumstances."*<br><br>*"But in reality, the social context can often be influenced to a limited extent, especially if the patient is socially isolated."* | Consensus was reached for this criterium.<br><br>From the comments we can conclude that the criterium was sufficiently clear. No exemptions or alternative wording options were proposed by respondents that can be expected to significantly alter the outcome in the following round.<br><br>We will therefore not repeat this criterium in the 2nd round and accept it as a consensus-criterium for irremediable |

| | | | |
|---|---|---|---|
| | | | psychiatric suffering. We will however describe these criteria in the executive summary. |
| 21. At least one recovery-oriented treatment must have been attempted without this leading to a significant reduction in suffering. | **Agree (72%)**<br><br>- Several respondents emphasize that all treatment should be recovery oriented.<br><br>- Others see recovery based (and patient oriented) treatment as especially important in the context of a PAD-request.<br><br>- Recovery based treatment should only be tried on indication. | *"Recovery focused approach is very important and often offers new perspectives."*<br><br>*"I think the concept of recovery is crucial in this issue"*<br><br>*"Every treatment trajectory should be focused on recovery"*<br><br>*"At least if the problem is such that there is an indication for Flexible assertive community treatment, or recovery-oriented treatment."* | Consensus was reached for this criterium.<br><br>From the comments we can conclude that the criterium was sufficiently clear. No exemptions or alternative wording options were proposed by respondents that can be expected to significantly alter the outcome in the following round.<br><br>We will therefore not repeat this criterium in the 2$^{nd}$ round and accept it as a consensus-criterium for irremediable psychiatric suffering. We will however describe these criteria in the executive summary. |

#### Dissensus criteria

| **Diagnostic criteria** |
|---|

| **Criterium** | **Respondents view on importance of the criterium** | **Example quotes** | **Steering group response** |
|---|---|---|---|
| 4. Broad psychodiagnostic testing, including personality testing, should be the standard, unless the psychiatrist provides clear reasons why it is not necessary. | **Disagree (36%)**<br>- Broad psychodiagnostics testing should not be standard, but only performed if indicated.<br>- No added value beyond a clinical diagnosis by two psychiatrists.<br>- Suffering is more important than a diagnosis.<br><br>**Agree (41%)**<br>- Diagnostic scrutiny is essential and the role broad psychodiagnostics testing can have in this.<br>- Recognizing a missed diagnosis may lead to missed treatment opportunities. | "That is not necessary by default, only on indication."<br><br>"I don't see the added value in that over a psychiatric examination by two psychiatrists."<br><br>"All suffering remains subjective and is an individual and unique experience of reality."<br><br>"I can't bear the thought that the diagnosis is wrong. That is one of the reasons that should be excluded in a euthanasia request (but preferably much earlier, of course). Because this can offer a different treatment perspective."<br><br>"I think it is very good to carry out extensive psychodiagnostic research and therefore also to conduct extensive personality research. It happens too often that patients do not recover from their initial diagnosis because personality problems are present but have not yet been diagnosed." | Consensus was not reached for this criterium.<br><br>We discussed this criterium and the comments in our steering group meeting. The words 'standard' and 'broad' are of little added value to this criterium and appear to make the criterium less clear to some of the respondents, which may influence the consensus-rate.<br><br>We therefore decided to change the criterium to: **Structured psycho-diagnostic testing, including personality testing when relevant, should be performed, unless the psychiatrist provides clear reasons why it is not necessary.**<br><br>In Dutch: **Psychodiagnostisch onderzoek moet worden ingezet, met inbegrip van persoonlijkheidsonderzoek wanneer relevant, tenzij de psychiater duidelijke redenen geeft waarom dit niet nodig is.** |

| | | | |
|---|---|---|---|
| 6. In addition to the descriptive diagnostics according to the DSM-5, a formulation must be drawn up for each patient based on a psychotherapeutic model relevant to the disorder. | **Disagree (30%)**<br>- A descriptive diagnosis is enough or even better.<br>- A PAD-procedure should not be seen as the start of a psychotherapeutic treatment.<br>- Specific psychotherapeutic models are of no added value.<br>- It should only be drawn up for specific disorders.<br><br>**Agree (43%)**<br>A psychotherapeutic formulation:<br>- Helps to better understand the suffering.<br>- emphasizes the importance of context.<br>- Is needed for a good diagnosis. | "In a descriptive diagnosis, the clinician tries to describe and explain both disorder and context as a solution direction. If that is done enough, a separate psychotherapeutic description is not necessary."<br><br>"Euthanasia is not a psychotherapeutic process. That may be the question in itself, but this is not the task of the independent expert to answer it."<br><br>"There is no single model that can reflect the often-complicated problems."<br><br>"With the more biologically oriented disorders such as schizophrenia and schizoaffective disorder, this is not always useful. This could be of added value if personality problems are present."<br><br>"Describing from a psychotherapeutic model helps to better understand the problem and also makes it clearer what has worked and what has not worked, or could work, based on this model."<br><br>"Yes, as long as there is an eye for the relational meaning of the wish to die."<br><br>"This too is self-evident to me and such a formulation belongs to adequate psychiatric diagnosis" | Consensus was not reached for this criterium.<br><br>In the steering group discussion, we acknowledged the comments. The use of the term 'based on a psychotherapeutic model relevant to the disorder' appears to lead to confusion and dissensus. This is not necessary. The essence of this criterium is to understand whether psychiatrists think that apart from the DSM-classification a more 'narrative diagnosis' is necessary when establishing irremediability.<br><br>Therefore, we changed the wording as following: '**When establishing irremediable psychiatric suffering a narrative account must be given, that includes etiology and pathogenesis, in addition to the classification according to the DSM-5.'**<br><br>In Dutch: '**Bij het vaststellen van niet-remedieerbaar psychiatrisch lijden moet, naast een DSM-classificatie, een beschrijvende diagnose gegeven worden met daarin een uitspraak over de etiologie en pathogenese'** |

| Treatment criteria | | | |
|---|---|---|---|
| **Criterium** | **Respondents view on importance of the criterium** | **Example quotes** | **Steering group response** |
| 12. When indicated, psychosurgical treatment (such as Deep Brain Stimulation) must have been attempted without significantly reducing suffering. | **Disagree (40%) / Agree (32%)**<br><br>**-** It should only be considered for certain indications.<br><br>- It is to experimental and does not have sufficient evidence base.<br><br>- It is to invasive or burdening for the patient.<br><br>- It should only be presented voluntarily. | "As far as I'm concerned, this only applies to patients with OCD (not patients with depression)."<br><br>“more invasive procedures (involving mortality) and experimental treatments should be considered optional”<br><br>“Demanding an invasive treatment with potentially serious side effects mandatory in order to qualify for assisted suicide is too high a threshold; you cannot demand this of anyone.”<br><br>“This is currently not a guideline-compliant treatment, so it cannot be required of the patient. It can be offered.” | Consensus was not reached for this criterium.<br><br>Again, the difference between suffering irremediably and establishing irremediable suffering was accepted.<br><br>The comments also implied that many respondents only viewed DBS as an ‘optional treatment’, we changed the wording reflecting this.<br><br>The new criterium is as follows: “**If indicated, psychosurgery (such as DBS) must be discussed and offered to the patient before irremediable psychiatric suffering can be established.”**<br><br>In Dutch: **wanneer psychochirurgie (zoals DBS) geïndiceerd is, moet dit eerst worden besproken en aangeboden, voordat niet-remedieerbaar psychiatrisch lijden vastgesteld kan worden.** |
| 16. If indicated, at least one acceptance-oriented psychotherapy must | **Disagree (9%) / Agree (60%)**<br><br>- It is seen by different respondents are an | *“In many cases, acceptance of psychological problems can lead to a reduction in the suffering of a patient.”* | *Consensus was not reached for this criterium.* |

| | | | |
|---|---|---|---|
| have been attempted without leading to a significant reduction in suffering before it can be considered irremediable. | important treatment step, especially for 'treatment-resistant' patients.<br><br>- Other respondents see practical difficulties i.e. availability of therapists.<br><br>- Treatment motivation is seen as crucial for the success of acceptance-based therapy, this may be lacking for patients requesting PAD. | *"In theory this is a nice statement, but in practice it is not always easy for someone to find a therapist who both masters the "correct" therapy, and who wants to accept the person in question as a patient and who has time for this."*<br><br>*"Once, provided the patient is open to it, otherwise in my opinion it has absolutely no effect to offer someone therapy if there is no clearly articulated and shared goal."* | In the steering group discussion, we argued that this criterium can be seen as confusing, due to the difference between suffering irremediably and establishing irremediable suffering. Therefore, we changed the criterium to: **If indicated, at least one acceptance-oriented psychotherapy must have been attempted without leading to a significant reduction in suffering before irremediable psychiatric suffering can be established.**<br><br>In Dutch: **Indien geïndiceerd, moet tenminste één op acceptatie gerichte psychotherapie zijn geprobeerd zonder dat dit heeft geleid tot een significante vermindering van het lijden voordat niet-remedieerbaar lijden vastgesteld kan worden.**<br>**(bijvoorbeeld: Acceptance and Commitment Therapy).** |
| 18. Indicated psychotherapeutic treatments that were ineffective in the past, should be repeated without leading to a significant reduction in suffering. | **Disagree (51%) / Agree (17%)**<br><br>**-** Only if the former psychotherapy was inadequate for some reason it should be repeated. | "Inquire how the previous therapy went and form a hypothesis as to why there was no effect. Depending on this, it may occasionally be considered to indicate a new treatment."<br><br>"People can also become 'treatment tired' and I think you have to pay attention to that. Another problem is that people are 'compulsory' to take a | Consensus was not reached for this criterium. We discussed the criterium with the steering group and based on the comments we propose the following reformulation for round 2:<br><br>'**If there are indications that entering into a repeated psychotherapeutic trajectory makes sense, a new trajectory must be offered before** |

| | | | |
|---|---|---|---|
| | - Only if the patient is motivated it should be repeated.<br>- Endless repetition of treatment offers little chance of success.<br>- Repetition of treatment should be at the psychiatrist's discretion. | therapy and then passively follow it and fail it, only to meet the criterion of euthanasia."<br>"This statement is gradually falling under 'therapeutic persistence'"<br>"This is at the discretion of the psychiatrist. If he/she is of the opinion that the psychotherapy has taken place well, this does not need to be repeated." | **irremediable psychiatric suffering can be established. For example: because conditions were sub-optimal in previous therapy'**<br>Dutch: **Als er aanwijzingen zijn dat het aangaan van een herhaald psychotherapeutisch traject zinvol is moet dit worden aangeboden, worden voordat niet-remedieerbaar psychiatrisch lijden vastgesteld kan worden. Bijvoorbeeld omdat de omstandigheden bij eerdere therapie suboptimaal waren.** |
| **Criteria concerning refusal** | | | |
| **Criterium** | **Respondents view on importance of the criterium** | **Example quotes** | **Steering group response** |
| 7. If a patient does not want to participate in the diagnostic process, there can be no irremediable psychiatric suffering. | **Disagree (26%) / Agree (49%)**<br>- The criterium is widely seen as internally inconsistent by both proponents and opponents: the suffering can be irremediable when a patient doesn't cooperate, but if a patient does not | *"There may still be an irremediable psychiatric condition, but it cannot be determined."*<br>*"Not wanting to cooperate with diagnosis can of course also be another symptom of a psychiatric illness"*<br>*"Just as you can't force someone to try every psychotropic drug, you can't force someone to take another test."* | None of the criteria concerning refusal reached consensus in round 1. We discussed these criteria in the steering group meeting and several conclusions can be drawn:<br>1. All refusal criteria led to confusion because it makes no distinction between suffering irremediably and establishing irremediability. This requires correction. |

| | | | |
|---|---|---|---|
| | cooperate it cannot be established as such.<br>- Non-compliance can be a symptom of (irremediable) psychiatric suffering.<br>- There should be a limit to the number of tests a patient is required to take. | | 2. Many respondents argue that there should be a 'limit' to the number of treatments or diagnostic procedures a patient should undergo.<br><br>2. We acknowledge from the comments that the respondents' considerations about refusal do not differ among treatments. Therefore, we choose not to repeat the changes for every different treatment in round 2.<br><br>3. Several respondents used the word 'reasonability' in relation to treatment refusal in their open definition and in the comments of the criteria. This was not accounted for in the round 1 criteria.<br><br>Based on the above we decided to comprise all treatment refusal criteria into the following two criteria:<br><br>A) **There are limits to the number of treatments a patient must undergo before it can be referred to as irremediable psychiatric suffering.**<br><br>**For example, patient or psychiatrist may refrain from further treatment on reasonable grounds, such as a long** |
| 9. When a patient refuses the abovementioned drug-treatments, the suffering is not irremediable. | **Disagree (23%) / Agree (53%)**<br><br>**-** Many respondents point out that the criterium is flawed: the suffering can be irremediable, but it cannot be established as such because the patient does not cooperate.<br><br>- There should be a reasonable limit to the amount of treatments a patient has to try. Different respondents view that at least the treatment protocol must be followed.<br><br>- The reason for treatment refusal is important and the | *"You cannot establish 'unremediable suffering' if the remedy has not been sufficiently attempted. Again, that doesn't mean it doesn't exist, but in a legal sense you can't use it."*<br><br>*"A person does not have to have been treated with every conceivable means. However, with what you can reasonably expect in the context of the guidelines and adequate medical action."*<br><br>*"That depends a lot on the reason for refusal (e.g. serious side effects in the patients history may be a reason not to try a certain treatment again)."*<br><br>*"I (..) can (…) imagine that patient and practitioner decide together to skip certain steps in a protocol or not to try them out. In short, joint decision-making is essential in this (…)."* | |

| | | | |
|---|---|---|---|
| | medication history plays a big role in establishing this.<br><br>- The decision about new treatment options should be based on shared decision making. | | **history of illness and treatment and / or the prospect of serious side effects.**<br><br>Dutch: **Er zijn grenzen aan de hoeveelheid behandelingen die een patiënt moet aangaan voordat er gesproken kan worden van niet remedieerbaar psychiatrisch lijden.**<br><br>**Bijvoorbeeld: patiënt of psychiater mogen afzien van verdere behandeling op redelijke gronden, zoals een lange geschiedenis van ziekte en behandeling en/of het vooruitzicht van ernstige bijwerkingen.**<br><br>B) **There are limits to the number of new diagnostic procedures a patient must undertake before it can be said that the psychiatric suffering is irremediable.**<br><br>**For example: a patient or psychiatrist may refrain from further treatment on reasonable grounds, such as a long history of illness and treatment.**<br><br>Dutch: **Er zijn grenzen aan de hoeveelheid nieuwe diagnostische procedures een patiënt moet aangaan voordat er gesproken kan worden van** |
| 11. When a patient refuses the abovementioned ECT, the suffering is not irremediable. | **Disagree (34%) / Agree (36%)**<br><br>**-** Again, different respondents point out that the suffering can be irremediable, but it cannot be established as such because the patient does not cooperate.<br><br>- Many respondents point to their earlier remarks, implying there is no substantial difference between refusing drug-treatment or ECT.<br><br>- ECT is not effective for ‘all suffering’.<br><br>- A patient has the right to refuse ECT and a doctor has the right to refuse PAD. | *“[Conform] supra, ECT is a proven efficacious treatment.”*<br><br>*“Then it cannot be determined, but it may indeed turn out to be irremediable.”*<br><br>*“ECT is not indicated for every psychiatric condition. It should be made clear that a condition for which ECT is indicated is thought to play a significant role in suffering.”*<br><br>*“Yet you also have the right to refuse treatment. Do you sometimes find that that person refuses an option that could very well reduce the psychological suffering, then you cannot take the path to euthanasia. At least - I wouldn't be able to.”*<br><br>*“Agreed, unless "forcing" someone to ECT actually increases suffering by side effects/feeling compelled.”* | |

| | | | |
|---|---|---|---|
| | - Not being able to refuse is coercive. | | **niet remedieerbaar psychiatrisch lijden.**<br><br>**Bijvoorbeeld: patiënt of psychiater mogen afzien van verdere diagnostiek op redelijke gronden, zoals een lange geschiedenis van ziekte en behandeling.** |
| 13. When a patient refuses the abovementioned psycho-surgical treatment, the suffering is not irremediable. | **Disagree (60%) / Agree (21%)**<br><br>- 23 respondents refer to their earlier argumentation (without further explanation).<br><br>- Psychosurgery is too invasive, burdensome and hazardous too demand.<br><br>- The evidence is insufficient.<br><br>- The suffering can be irremediable, but not established as such. | *"Same explanation as with ECT."*<br><br>*"One can never force people into (invasive/experimental) treatment, let alone if it cannot be predicted with certainty that it would provide a solution to unbearable suffering."*<br><br>*"Difficult. Can you oblige a patient to undergo DBS before he can be euthanized? I think that the developments of DBS are not yet far enough for that."*<br><br>*"I'm still only talking about patients with OCD. Practically speaking, I agree. But strictly speaking, you can't say that someone's suffering cannot be cured if you haven't actually done the best you can, because you simply don't know that. There could be irremediable suffering, but you have not been able to determine that. The suffering MAY be curable because not everything has been tried."* | |
| 15. When a patient refuses the abovementioned psychotherapy, the suffering is not irremediable. | **Disagree (17%) / Agree (57%)**<br><br>- 13 respondents refer to their earlier argumentation (without further explanation).<br><br>- Respondents, again, point out that the suffering can be | *"Again it could be, but I can't determine it."*<br><br>*"Psychotherapy can make an essential difference in dealing with suffering in many psychiatric disorders, so even if it cannot make a curative difference, the use of psychotherapy is essential."*<br><br>*"Situations can be envisaged in which, for example, history, it can be concluded that further* | |

| | | |
|---|---|---|
| | irremediable, but it cannot be established as such because the patient does not cooperate.<br><br>- Psychotherapy is seen as an important standard treatment.<br><br>- Respondents give many different exemptions for which treatment refusal should be accepted.<br><br>- Treatment motivation is an important factor in psychotherapy, both among supporters and opponents of this criterion (see examples). | *psychotherapeutic treatments no longer have any added value."*<br><br>*"If I think that in principle the patient can cooperate with the treatment and after dependent resistance a treatment can still be started, then I will demand that."*<br><br>*"Psychotherapy against the will of the patient cannot succeed. Needing things can be part of suffering."* |
| 17. When a patient refuses the abovementioned acceptance-oriented psychotherapy, the suffering is not irremediable. | **Disagree (23%) / Agree (47%)**<br><br>- 15 respondents refer to their earlier argumentation (without further explanation).<br><br>- Respondents, again, point out that the suffering can be irremediable, but it cannot be established as such | *"Again, I think the question is incorrect. If someone does not have something investigated, then no conclusion can be drawn."*<br><br>*"That's right, you are missing an important treatment that could make you better or suffer less from your complaints."*<br><br>*"Depends on the guideline and how many other therapies have already been done."* |

| | | |
|---|---|---|
| | because the patient does not cooperate.<br><br>- Acceptance-oriented psychotherapy is an important treatment.<br><br>- Different adjuvant conditions for the criterion are given, for instance: it depends on the treatment history, it is always individual or it depends on the reason for refusal. | *"This depends on whether a consensus can be reached in the dialogue about the unbearableness and the hopelessness."* |
| 19. When a patient refuses the abovementioned repetition of psychotherapy, the suffering is not irremediable. | **Disagree (47%) / Agree (11%)**<br><br>- 13 respondents refer to their earlier argumentation (without further explanation).<br><br>- Whether this should be a criterion is said to depend on the quality of the earlier treatment.<br><br>- Other respondents argue that this also depends on patient factors. | *"A patient may refuse a previously pointless treatment. Incidentally, it can of course be checked whether it has really been an adequate psychotherapy by a recognized psychotherapist. Therapy by a basic psychologist is of course different from psychotherapy by a clinical psychologist/psychiatrist, for example."*<br><br>*"Depending on the explanation why it didn't work before. For example, low intelligence? Other patient-related characteristics that interfere with psychotherapy and chance of success. E.g. antisocial characteristics?"* |

### Hyperlink to complete results (in Dutch)

Click here to open file.
