## Supplement 2 for "Irremediable psychiatric suffering in the context of physician assisted death: a Delphi-study"

### Irremediable psychiatric suffering - Delphi round 2 - Feedback report

#### Table of contents

Table of contents 1
Executive summary 2
Quantitative data summary 3
Response rate 3
Consensus criteria 3
Dissensus criteria 4
Qualitative content analysis of respondent comments 5
Consensus criteria 5
Dissensus criteria 9
Summary of consensus-criteria from round 1 and 2 13
Diagnostic criteria 13
Treatment criteria 13
Hyperlink to complete results (Dutch) 14

### Executive summary

The aim of this Delphi-study is to find expert-consensus on the concept of irremediable psychiatric suffering (IPS) in the context of physician assisted death (PAD). In round 1 we included 53 psychiatrists who have experience with PAD from the Netherlands and Belgium. They gave their own definition of IPS and gave their views on potential criteria for IPS in the context of PAD through a Likert scale and open comments.

After round 1 the open definition and criteria were analyzed and discussed in 2 steering group meetings. The open definitions led us to 2 additional criteria; one concerning a demand for the duration of suffering and one introducing a retrospective definition of IPS, based on failed treatments. Of the 21 potential criteria from round 1 the Likert-scales were calculated and the comments were analyzed through (qualitative) direct content analysis. All results were subsequently discussed in the steering group. 8 criteria, all concerning treatment and diagnosis, were accepted as consensus-criteria. 13 criteria were recognized as dissensus-criteria and subsequently rephrased or restructured for round 2. For a more detailed description of these criteria and the design of round 2, please find the complete feedback report of round 1 on the open science framework (project code: qx5hy).

Eventually, after two steering group discussions, 9 potential criteria were resubmitted to the 53 respondents in round 2. 47 psychiatrists responded in this round (89% response rate), and the criteria and comments were again analyzed using Likert-scales and direct content analysis. This time 5 out of 9 criteria reached consensus and 4 criteria did not. Through the qualitative analysis of the comments we could conclude that the questions were sufficiently clear and the respondents consequently applied their arguments for agreeing or disagreeing, we therefore accepted all criteria as either consensus or dissensus criteria and saw no added value of a third round.

The consensus criteria in round 2 were the following. First, because all reasonable treatments must be tried, psychiatric suffering must be present for several years before IPS can be established. Second, when establishing irremediable psychiatric suffering a narrative account must be given, that includes etiology and pathogenesis, in addition to the classification according to the DSM-5. Third, if there are indications that entering into a repeated psychotherapeutic trajectory makes sense, a new trajectory must be offered before irremediable psychiatric suffering can be established. Fourth, there are limits to the number of treatments a patient must undergo before it can be referred to as irremediable psychiatric suffering. Fifth, there are limits to the number of new diagnostic procedures a patient must undertake before it can be said that the psychiatric suffering is irremediable.

The dissensus criteria in round 2 were the following. First, because it is often difficult to establish a reliable prognosis, the judgment about non-remediable psychiatric suffering must be based on the failure of treatment in the past. Second, structured psycho-diagnostic testing, including personality testing when relevant, should be performed, unless the psychiatrist provides clear reasons why it is not necessary. Third, if indicated, psychosurgery (such as DBS) must be discussed and offered to the patient before irremediable psychiatric suffering can be established. Fourth, if indicated, at least one acceptance-oriented psychotherapy must have been attempted without leading to a significant reduction in suffering before irremediable psychiatric suffering can be established.

### Quantitative data summary

#### Response rate

47 of 53 (89%) psychiatrists who responded to the first round also responded to this second round.

#### Consensus criteria

| Round 2 criteria | % agree or strongly agree |
|---|---|
| 2. Because all reasonable treatments must be tried, the psychiatric suffering must be present for several years before irremediable psychiatric suffering can be established. | **81** |
| 4. When establishing irremediable psychiatric suffering a narrative account must be given, that includes etiology and pathogenesis, in addition to the classification according to the DSM-5. | **91** |
| 7. If there are indications that entering into a repeated psychotherapeutic trajectory is meaningful, this must be offered before irremediable psychiatric suffering can be established.<br>*For example: because conditions were sub-optimal in previous therapy.* | **70** |
| 8. There are limits to the number of treatments a patient must undergo before it can be referred to as irremediable psychiatric suffering.<br>*For example, patient or psychiatrist may refrain from further treatment on reasonable grounds, such as a long history of illness and treatment and / or the prospect of serious side effects.* | **81** |
| 9. There are limits to the number of new diagnostic procedures a patient must undertake before it can be said that the psychiatric suffering is irremediable.<br>*For example: a patient or psychiatrist may refrain from further treatment on reasonable grounds, such as a long history of illness and treatment.* | **81** |

#### Dissensus criteria

| Round 2 criteria | % disagree or strongly disagree | % agree or strongly agree |
|---|---|---|
| 1. Because it is often difficult to establish a reliable prognosis, the judgment about non-remediable psychiatric suffering must be based on the failure of treatment in the past. | **11** | **66** |
| 3. Structured psycho-diagnostic testing, including personality testing when relevant, should be performed, unless the psychiatrist provides clear reasons why it is not necessary. | **32** | **55** |
| 5. If indicated, psychosurgery (such as DBS) must be discussed and offered to the patient before irremediable psychiatric suffering can be established. | **28** | **62** |
| 6. If indicated, at least one acceptance-oriented psychotherapy must have been attempted without leading to a significant reduction in suffering before irremediable psychiatric suffering can be established. | **13** | **66** |

### Qualitative content analysis of respondent comments

#### Consensus criteria

Consensus was defined that 70% or more of the respondents agreed or strongly agreed, or inversely disagreed or strongly disagreed with a criterium. Five of nine criteria reached consensus in the first round.

| Diagnostic criteria | | | |
|---|---|---|---|
| **Criterium**<br><br>**+ Relation of criterium to round 1.** | **Respondents view on importance of the criterium** | **Example quotes** | **Steering group response** |
| 2. Because all reasonable treatments must be tried, the psychiatric suffering must be present for several years before non-remedial measures can be established.<br><br>**Relation to round 1:** this is a newly added criterium, based on the open definition of IPS the respondents gave in round 1. | There was a broad consensus (81%) for this criterium. 74% of respondents commented on their answer.<br><br>Different respondents argue that psychiatric treatment takes time in practice, therefore they agree with the criterium.<br><br>Time is also seen as an important prognostic factor in itself.<br><br>Another often mentioned comment is that there are exceptions to this criterium. | *"After all, it takes time to try all reasonable treatments! I cannot imagine a psychiatric disorder, ie, of which it is clear from the outset that the prognosis is very poor (except for neuropsychiatric problems, eg in M. Huntington)."*<br><br>*"Time can also contribute to context change and personal growth."* | **Group discussion:** from the comments we can conclude that the criterium was sufficiently clear. No exemptions or alternative wording options were proposed by respondents that can be expected to significantly alter the outcome in the following round.<br><br>**Decision:** we will therefore not repeat this criterium in a next round and accept it as a consensus-criterium for irremediable psychiatric suffering. |

| | | | |
|---|---|---|---|
| 4. When establishing irremediable psychiatric suffering a narrative account must be given, that includes etiology and pathogenesis, in addition to the classification according to the DSM-5.<br><br>**Relation to round 1:** this question is an adjustment to the criterium in round one stating *'In addition to the descriptive diagnostics according to the DSM-5, a formulation must be drawn up for each patient based on a psychotherapeutic model relevant to the disorder.'* It did not reach consensus (30% disagreed and 43% agreed). From the comments we learned that many respondents had trouble with the term 'based on a psychotherapeutic model' so we removed this and discussed that the essence of this criterium, in our view, is a 'narrative' approach to psychiatric diagnostics, so we added this description in the round 2 criterium. | There was a broad consensus (91%) for this criterium. 70% of respondents commented on their answer.<br><br>Several respondents comment that the narrative diagnosis is an important element of psychiatric practice. Some go on to say that the DSM is not important.<br><br>Different respondents agree with the removal of 'psychotherapeutic model'. | *"A narrative diagnosis in this vein is certainly part of it."*<br><br>*"the parenthesis 'next to a DSM classification' could even be removed, a descriptive diagnosis is more important (in any context, in my view)"*<br><br>*"It is good that the psychotherapeutic model has been removed from the question"* | **Group discussion:** this criterium adds nuance to the DSM5, recognizing that not only a diagnostic classification is important. Prof. Widdershoven pointed out that in somatic euthanasia cases the 'story' (e.g. from the GP) would also be important.<br><br>From the comments we can conclude that the criterium was sufficiently clear. No exemptions or alternative wording options were proposed by respondents that can be expected to significantly alter the outcome in the following round.<br><br>**Decision:** we will therefore not repeat this criterium in a next round and accept it as a consensus-criterium for irremediable psychiatric suffering. |
| 7. If there are indications that entering into a repeated psychotherapeutic trajectory is meaningful, this must be offered before irremediable psychiatric suffering can be established. | There was a narrow consensus (70%) for this criterium. 55% of respondents commented on their answer. | *"If there are indications that this could be useful, then it should be tried. After all, death is final."*<br><br>*"Depends on the reason the therapy didn't help, what* | **Group discussion:** based on the comments one can argue that different interpretations of this criterium were possible, further specification of the criterium might |

| | | | |
|---|---|---|---|
| **Relation to round 1:** this question is an adjustment to the criterium in round one stating *'indicated psychotherapeutic treatments that were ineffective in the past, should be repeated without leading to a significant reduction in suffering.'* It did not reach consensus (26% disagreed and 49% agreed). From the comments we learned that different respondents only saw value of repeating 'inadequately performed' psychotherapy. Also, different respondents argued that repeated therapy should only be *offered,* not *demanded*. We adjusted the criterium to reflect these comments. | Several respondents merely commented that they agreed with this statement.<br><br>Others stated that the nature of the past psychotherapy is an important factor in their view on this criterium.<br><br>It was also mentioned that this criterium is only applicable to motivated patients. | *conditions were sub-optimal, and is this different now?"*<br><br>*"It can be offered, but if the patient isn't motivated for it, it won't yield any benefits."* | be possible, but in our view the criterium was sufficiently clear.<br><br>Also, addition of a separate criterium to emphasize the importance of repeated pharmacological interventions, was discussed, but the idea was eventually discarded. First because the need for adequate pharmacological treatment is already recorded as a consensus criterium in round 1, repetition of a inadequate treatment is part of that, this is less clear for psychotherapy. Secondly prof. Beekman mentioned in the discussion that in his view this criterium rightly emphasizes the importance of thorough psychotherapy.<br><br>**Decision:** based on the abovementioned considerations we will not repeat this criterium in a next round or add new criteria based on this criteriums comments. We accept this consensus criterium for irremediable psychiatric suffering. |

| | | | |
|---|---|---|---|
| 8. There are limits to the number of new diagnostic procedures a patient must undertake before it can be said that the psychiatric suffering is irremediable.<br>For example: a patient or psychiatrist may refrain from further treatment on reasonable grounds, such as a long history of illness and treatment.<br><br>**Relation to round 1:** in round 1 we (separately) asked whether patients could refuse a broad range of treatments and diagnosis. All of these criteria did not reach consensus. The respondents commented that there is a difference between establishing irremediability and suffering irremediable, a distinction that was not accounted for in round 1. They also commented that treatment refusal can be reason not to establish IPS, but that there should be *limits.* And finally, they commented that their views on treatment refusal were the same for different treatments. These comments were combined in question 8 and 9 in round 2. | There was a broad consensus (81%) for this criterium. 47% of respondents commented on their answer.<br><br>Several respondents comment that they agree with the criterium.<br><br>Others argue that the decision-making process should be left to the individual psychiatrist and patient. | *"Good definition"*<br><br>*"What are reasonable grounds? The psychiatrist must estimate whether further diagnostics are useful, you cannot capture that in a guideline."* | **Group discussion:** in the group discussion dr. Evans agreed with from the questions about the nature of reasonability and suggested we should add this in our discussion. Furthermore it was concluded in the meeting that the criterium was sufficiently clear. No exemptions or alternative wording options were proposed by respondents that can be expected to significantly alter the outcome in the following round.<br><br>**Decision:** we will therefore not repeat this criterium in a next round and accept it as a consensus-criterium for irremediable psychiatric suffering. |
| 9. There are limits to the number of treatments a patient must undergo before it can be referred to as irremediable psychiatric suffering.<br>For example, patient or psychiatrist may refrain from further treatment on reasonable grounds, such as a long history of illness and treatment and / or the prospect of serious side effects. | There was a broad consensus (81%) for this criterium. 45% of respondents commented on their answer. | *"No, if the treatment can alleviate the suffering, then that treatment should not be waived. and: what is the prospect of serious side effects? ECT, for example, should not be refused because of "the prospect of serious side effects". If the patient improves thanks* | **Group discussion:** from the comments we can conclude that the criterium was sufficiently clear. No exemptions or alternative wording options were proposed by respondents that can be expected to significantly alter the outcome in the following round. |

| | | | |
|---|---|---|---|
| **Relation to round 1:** in round 1 we (separately) asked whether patients could refuse a broad range of treatments and diagnosis. All of these criteria did not reach consensus. The respondents commented that there is a difference between establishing irremediability and suffering irremediable, a distinction that was not accounted for in round 1. They also commented that treatment refusal can be reason not to establish IPS, but that there should be *limits.* And finally, they commented that their views on treatment refusal were the same for different treatments. These comments were combined in question 8 and 9 in round 2. | Different respondents refer to their comment for criterium 8.<br><br>Some respondents argue that all real treatment options have to be tried.<br><br>Other respondents argue that the length and quality of earlier treatment is important when evaluating this criterium. | *to the ECT and then finds that the memory complaints are so serious that he does not want to live, then we will talk further."*<br><br>*"That very much depends on whether the previous treatments have been carried out adequately."* | **Decision:** we will therefore not repeat this criterium in a next round and accept it as a consensus-criterium for irremediable psychiatric suffering. |

#### Dissensus criteria

| **Diagnostic criteria** | | | |
|---|---|---|---|
| **Criterium**<br><br>**+ Explanation about relation of criterium to round 1.** | **Respondents view on importance of the criterium** | **Example quotes** | **Steering group response** |
| **1. Because it is often difficult to establish a reliable prognosis, the judgment about non-remediable psychiatric suffering must be based on the failure of treatment in the past.** | This criterium did not reach consensus (11% disagreed and 66% agreed). 83% of respondents commented on their answer.<br><br>Several psychiatrists argued that a purely retrospective | *"Agree, but not just based on this; other prognostic factors should be taken into account."*<br><br>*"It does give more information, but it doesn't fully cover the load."* | **Group discussion:** from the responses we conclude that the question was sufficiently clear. Although consensus was almost reached, different areas of disagreement remain as described on the left. |

| | | | |
|---|---|---|---|
| **Relation to round 1:** this is a newly added criterium, based on the open definition of IPS the respondents gave in round 1. | view on IPS is to reductive. The psychiatrist must also form an opinion about the prognosis, even if this is complicated.<br><br>Other psychiatrists emphasize the importance of the quality of earlier treatments. | *"Of course, a thorough evaluation must be made of whether the previous treatment has been adequately carried out in terms of technical aspects and treatment."* | **Decision:** we accept that this is a dissensus criterium and will not repeat this criterium in a new round. |
| **3. Structured psycho-diagnostic testing, including personality testing when relevant, should be performed, unless the psychiatrist provides clear reasons why it is not necessary.**<br><br>**Relation to round 1:** in round one the criterium was '*Broad psychodiagnostic testing, including personality testing, should be the standard, unless the psychiatrist provides clear reasons why it is not necessary.'* This did not reach consensus (36% disagreed / 41% agreed). We changed the wording because several respondents appeared to have trouble with the word 'broad' or 'standard', which we deemed of little added value. | This criterium did not reach consensus (32% disagreed and 55% agreed). 77% of respondents commented on their answer.<br><br>Several respondents argue that it should only be performed if indicated.<br><br>A division can be seen in the comments between psychiatrists who see added value of structured psychodiagnostics and those who don't. | *"I can agree with that, but I would not oblige it, but strongly recommend it."*<br><br>*"Yes, it provides insight into the dynamics and can provide explanations for why there is no recovery and the person is stuck and has a death wish. It is better to determine whether it is hopeless or not."*<br><br>*"I think a good history, biography, an interview with important peers and a psychiatric examination say more than psychodiagnostic tests."* | **Group discussion:** from this round we can conclude that this word change, or reading the arguments from other respondents, did not substantially change the views on a group level. Prof. Beekman pointed out that it is an established fact that psychiatrists tend to overestimate their diagnostic and prognostic abilities, this may have skewed their views on this criterium.<br><br>**Decision:** we therefore accept that this is a dissensus criterium and will not repeat this criterium in a new round. |

| | | | |
|---|---|---|---|
| **5. If indicated, psychosurgery (such as DBS) must be discussed and offered to the patient before irremediable psychiatric suffering can be established.**<br><br>**Relation to round 1:** in round 1 the criterium was *'When indicated, psychosurgical treatment (such as Deep Brain Stimulation) must have been attempted without significantly reducing suffering.'* This did not reach consensus (40% disagreed and 32% agreed). The comments in round 1 also showed that many respondents thought psychosurgery should be discussed but not mandated, so we changed the wording accordingly. | This criterium did not reach consensus (28% disagreed and 62% agreed). 70% of respondents commented on their answer.<br><br>Several respondents mentioned that psychosurgery should be discussed, but not mandated.<br><br>Other respondents commented on the (perceived) experimental nature of DBS and the invasiveness. | *"I believe it should be discussed, if indicated. In my view, the patient may refuse this"*<br><br>*"I find these treatment methods experimental."*<br><br>*"Surgery is always an invasive procedure, so the possible result is less related to the patient's autonomy and will."* | **Group discussion:** changing the wording did almost lead to consensus, but still a substantial portion of respondents disagreed, mainly due to the invasiveness and (perceived) experimental nature of psychosurgery. The comments did not show that the authors misunderstood or misinterpreted the criterium.<br><br>**Decision:** we accept that this is a dissensus criterium and will not repeat this criterium in a new round. |
| **6. If indicated, at least one acceptance-oriented psychotherapy must have been attempted without leading to a significant reduction in suffering before irremediable psychiatric suffering can be established.**<br><br>**Relation to round 1:** In round 1 the criterium was "*If indicated, at least one acceptance-oriented psychotherapy must have been attempted without leading to a significant reduction in suffering before it can be considered irremediable."* This did not reach consensus (9% disagreed and | This criterium did not reach consensus (13% disagreed and 66% agreed). 45% of respondents commented on their answer.<br><br>Different respondents mention the importance of acceptance-based treatment.<br><br>However, motivation is seen as an important requirement | *"Dealing better with complaints and accepting limitations can lead to more joy in life and less suffering, and as such should be seen as a potentially effective treatment that should be tried out."*<br><br>*"I don't think it makes sense to regulate everything in advance. With the guidelines in hand, the psychiatrist must come to the conclusion that the disorder is* | **Group discussion:** the change to the criterium and showing the psychiatrists their peers comments did not change the outcome, dissensus continued. The comments did not show that the authors misunderstood or misinterpreted the criterium.<br><br>Based on the comments dr. Evans pointed out that it may be possible that the views on some criteria are dependent on the psychiatric |

| | | | |
|---|---|---|---|
| 60% agreed). The comments mainly showed that the respondents saw a difference between 'suffering irremediable' and 'establishing irremediable suffering'. We changed the criterium accordingly and repeated it in round 2 with comments from round 1. | and it should be limited in time.<br><br>Other respondents did not see the specific merit of acceptance-based psychotherapy or did not see the merit of drawing up specific criteria. | *treatment-resistant with no prospect of improvement."*<br><br>*"The therapy must be supported [by the patient], there should be a good therapeutic relationship and the duration of therapy must be clearly defined."* | 'school' a respondent/expert adheres to, this may mean that certain criteria are suited for different areas of psychiatry where a particular school is more relevant.<br><br>**Decision:** we accept that this is a dissensus criterium and will not repeat this criterium in a new round. |

### Summary of consensus-criteria from round 1 and 2

#### Diagnostic criteria

A. When establishing irremediable psychiatric suffering:
   1. A psychiatric diagnosis, as described in the DSM-5, should be established according to applicable guidelines.
   2. In addition to the classification according to the DSM-5, a narrative account must be given that includes etiology and pathogenesis.
   3. In addition to the descriptive diagnostics according to the DSM-5, it should be standard practice to verify whether there are contextual or systemic factors that cause or maintain the psychiatric complaints.

B. During the PAD-procedure, the diagnosis must be independently confirmed by at least two psychiatrists.

C. There are limits to the number of new diagnostic procedures a patient must undertake before it can be said that the psychiatric suffering is irremediable. *For example: a patient or psychiatrist may refrain from further diagnostic procedures on reasonable grounds, such as a long history of illness and treatment.*

#### Treatment criteria

D. If side effects allowed, the indicated drug-treatments should have been adequately performed without leading to a significant reduction in suffering.

E. If side effects allowed and if indicated, electroconvulsive therapy (ECT) should have been attempted for a sufficient length of time without leading to a significant reduction in suffering.

F. Psychotherapeutic treatments indicated by the applicable guideline must have been attempted without leading to a significant reduction in suffering.

G. If there are indications that entering into a repeated psychotherapeutic trajectory is meaningful, this must be offered before irremediable psychiatric suffering can be established. *For example: because conditions were sub-optimal in previous therapy.*

H. At least one recovery-oriented treatment must have been attempted without leading to a significant reduction in suffering.

I. If necessary, substantial efforts should have been made to improve the patient's social situation without leading to a significant reduction in suffering.

J. Because all reasonable treatments must be tried, the psychiatric suffering must have been present for several years before irremediable psychiatric suffering can be established.

K. There are limits to the number of treatments a patient must undergo before psychiatric suffering can be considered irremediable. *For example, a patient or psychiatrist may refrain from further treatment on reasonable grounds, such as a long history of illness and treatment or the prospect of serious side effects.*

#### Hyperlink to complete results (in Dutch)

Click here to open file.
